# Impact of Delta on viral burden and vaccine effectiveness against new SARS-CoV-2 infections in the UK

**DOI:** 10.1101/2021.08.18.21262237

**Authors:** Koen B. Pouwels, Emma Pritchard, Philippa C. Matthews, Nicole Stoesser, David W. Eyre, Karina-Doris Vihta, Thomas House, Jodie Hay, John I Bell, John N Newton, Jeremy Farrar, Derrick Crook, Duncan Cook, Emma Rourke, Ruth Studley, Tim Peto, Ian Diamond, A. Sarah Walker, the COVID-19 Infection Survey Team

## Abstract

The effectiveness of BNT162b2, ChAdOx1, and mRNA-1273 vaccines against new SARS-CoV-2 infections requires continuous re-evaluation, given the increasingly dominant Delta variant. We investigated the effectiveness of the vaccines in a large community-based survey of randomly selected households across the UK. We found that the effectiveness of BNT162b2 and ChAd0x1 against any infections (new PCR positives) and infections with symptoms or high viral burden is reduced with the Delta variant. A single dose of the mRNA-1273 vaccine had similar or greater effectiveness compared to a single dose of BNT162b2 or ChAdOx1. Effectiveness of two doses remains at least as great as protection afforded by prior natural infection. The dynamics of immunity following second doses differed significantly between BNT162b2 and ChAdOx1, with greater initial effectiveness against new PCR-positives but faster declines in protection against high viral burden and symptomatic infection with BNT162b2. There was no evidence that effectiveness varied by dosing interval, but protection was higher among those vaccinated following a prior infection and younger adults. With Delta, infections occurring following two vaccinations had similar peak viral burden to those in unvaccinated individuals. SARS-CoV-2 vaccination still reduces new infections, but effectiveness and attenuation of peak viral burden are reduced with Delta.

## Introduction

Multiple studies have assessed the real-world effectiveness of different COVID-19 vaccination programs in the general population, healthcare and other frontline workers, and care home residents^1^. Studies generally showed high effectiveness of the BNT162b2 mRNA vaccine (Pfizer-BioNTech) and the Oxford-AstraZeneca adenovirus-vector vaccine, ChAdOx1 nCoV-19 (termed here ChAdOx1) against the Alpha (B.1.1.7) and preceding variants. More limited real-world effectiveness data is available for the mRNA-1273 (Moderna) vaccine^2-4^. Continued emergence of new SARS-CoV-2 variants potentially threatens the success of vaccination programs, particularly as *in vitro* experiments suggest reduced neutralisation activity of vaccine-elicited antibodies against emerging variants^5,6^. Of particular concern is the Delta variant (B.1.617.2), which has caused sharp rises in infections in many countries, including some with relatively high vaccination coverage such as the UK. In England, Delta quickly became dominant after being classified as a Variant of Concern on 28 April 2021, reaching 61% of sequenced positives from the English symptomatic testing program in the week commencing 17 May^7^ and 99% from 27 June onwards^8^.

Real-world data on vaccine effectiveness against Delta infections are currently limited. A test-negative case-control study using data to 16 May 2021 from the English symptomatic testing program suggested that the effectiveness after one BNT162b2 or ChAdOx1 vaccination was lower against symptomatic infection with Delta (31%) than Alpha (49%)^9^. Reductions in effectiveness against infection with Delta versus Alpha were smaller following two doses of either vaccine. However, estimates from test-negative case-control studies may be biased if vaccination status influences test-seeking behaviour of cases not requiring healthcare^10^. A recent study from Scotland also suggested reduced effectiveness against infection with Delta versus Alpha following two doses of either vaccine^11^. However, they found no evidence that effectiveness on hospital admissions among those first testing positive varied with Delta versus Alpha, leaving it unclear to what extent the results for infection might be attributable to bias due to test-seeking behaviour being influenced by vaccination status^10^. A further contributor may be waning immunity, with two recent studies from Israel finding higher infection rates in those vaccinated earliest^12,13^.

We therefore assessed the effectiveness of the BNT162b2, ChAdOx1, and mRNA-1273 vaccines against new SARS-CoV-2 PCR-positive tests using the Office for National Statistics COVID-19 Infection Survey, a large community-based survey of individuals living in randomly selected private households across the UK, where RT-PCR tests were performed following a pre-determined schedule, irrespective of symptoms, vaccination and prior infection^14,15^. We assessed vaccine effectiveness based on overall RT-PCR positivity, and split according to self-reported symptoms, cycle threshold (Ct) value (<30 versus ≥30) as a surrogate for viral load, from 1 December 2020 (start of vaccination rollout) to 16 May 2021, when Alpha dominated, and from 17 May 2021 to 1 August 2021 when Alpha was replaced by Delta (**Figure S1**; confirmed by available whole genome sequencing^16^). In addition, in this Delta-dominant period, we investigated variation in vaccine effectiveness by long-term health conditions, age (18-34 versus 35-65 years) interval between first and second vaccination (<9 weeks versus ≥9 weeks), and prior infection. We also assessed viral burden in new PCR-positives occurring ≥14 days after second vaccination using Ct values.

## Results

### Visits and new PCR-positives included in analysis

During the Alpha-dominant period from 1 December 2020 to 16 May 2021 (**Figure S1**), nose and throat RT-PCR results were obtained from 384,543 individuals aged 18 years or older (221,909 households) at 2,580,021 visits (median [IQR] 7 [6-8]), of which 16,538 (0.6%) were the first PCR-positive in a new infection episode. During the Delta-dominant period from 17 May to 1 August 2021, results were obtained from 358,983 individuals (213,825 households) at 811,624 visits (median [IQR] 2 [2-3]), 3,123 (0.4%) being the first PCR-positive. Characteristics at included visits are shown in **Table S1**.

We classified each visit according to vaccination status and previous infection as previously^15^ (**Table S2**), considering those not yet vaccinated or >21 days before vaccination without evidence of prior infection as the reference group. The vast majority of post-vaccination visits between 1 December 2020 and 16 May 2021 were from individuals that received BNT162b2 or ChAdOx1, but from 17 May 2021 there were also visits from individuals that received mRNA-1273. The median (IQR) time since first vaccination for visits ≥21 days after the first vaccination but before the second was 47 (34-61), 43 (31-58), and 41 (31-52) for ChAdOx1, BNT162b2 and mRNA-1273, respectively; and from second vaccination for visits ≥14 days after the second vaccination 41 (27-57) and 59 (35-86), respectively (insufficient data for mRNA-1273). The median (IQR) dosing interval between first and second vaccination was 76 (68-78) days for ChAdOx1 and 74 (62-77) for BNT162b2.

### Impact of vaccination on new PCR-positives, regardless of self-reported symptoms

Adjusting for multiple potential confounders, in the Alpha-dominant period the vaccine effectiveness (VE) of both BNT162b2 and ChAdOx1 vaccines against new PCR-positives was similar amongst those ≥18 years to that previously reported to 8 May 2021 amongst those ≥16 years^15^ (**Table 1**).

**Table 1.**
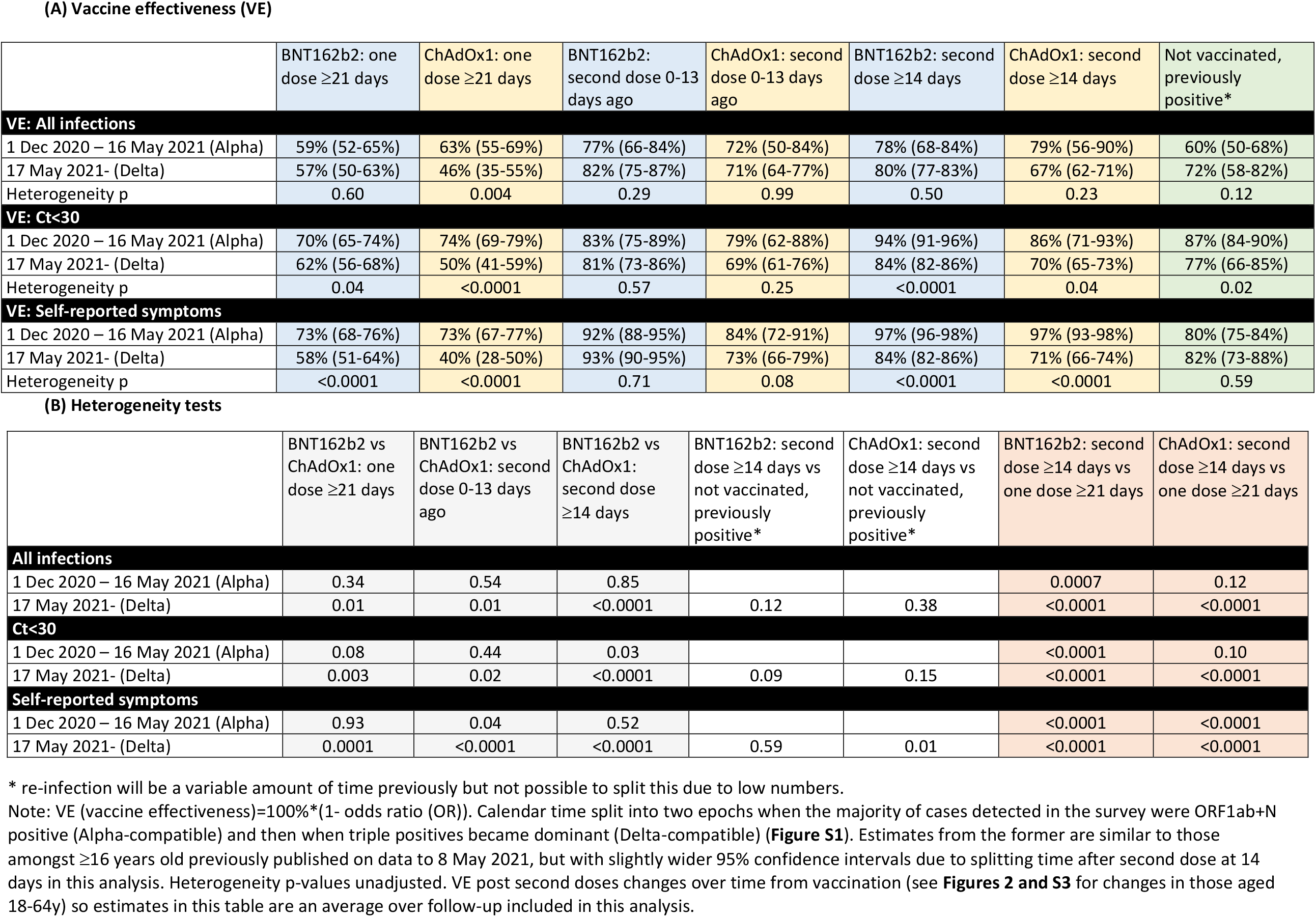
**Vaccine effectiveness (95% CI) (A) and comparisons between vaccines and with previous infection (B) in those aged 18 years and older in Alpha-dominant and Delta-dominant periods**

In the Delta-dominant period, amongst those ≥18 years there was evidence of reduced effectiveness ≥21 days after the first ChAdOx1 vaccination (VE 46% (95% CI 35-55%), heterogeneity p=0.004), but not ≥14 days after the second (67%, 62-71% vs 79%, 56-90% in the Alpha-dominant period, heterogeneity p=0.23). There was no evidence of reduced effectiveness in the Delta-dominant period for BNT162b2 against all new PCR-positives, with VE 57% (50-63%) post first dose and 80% (77-83%) post second dose (heterogeneity p=0.60, p=0.23, respectively) (**Table 1, Figure 1**).

**Figure 1.**
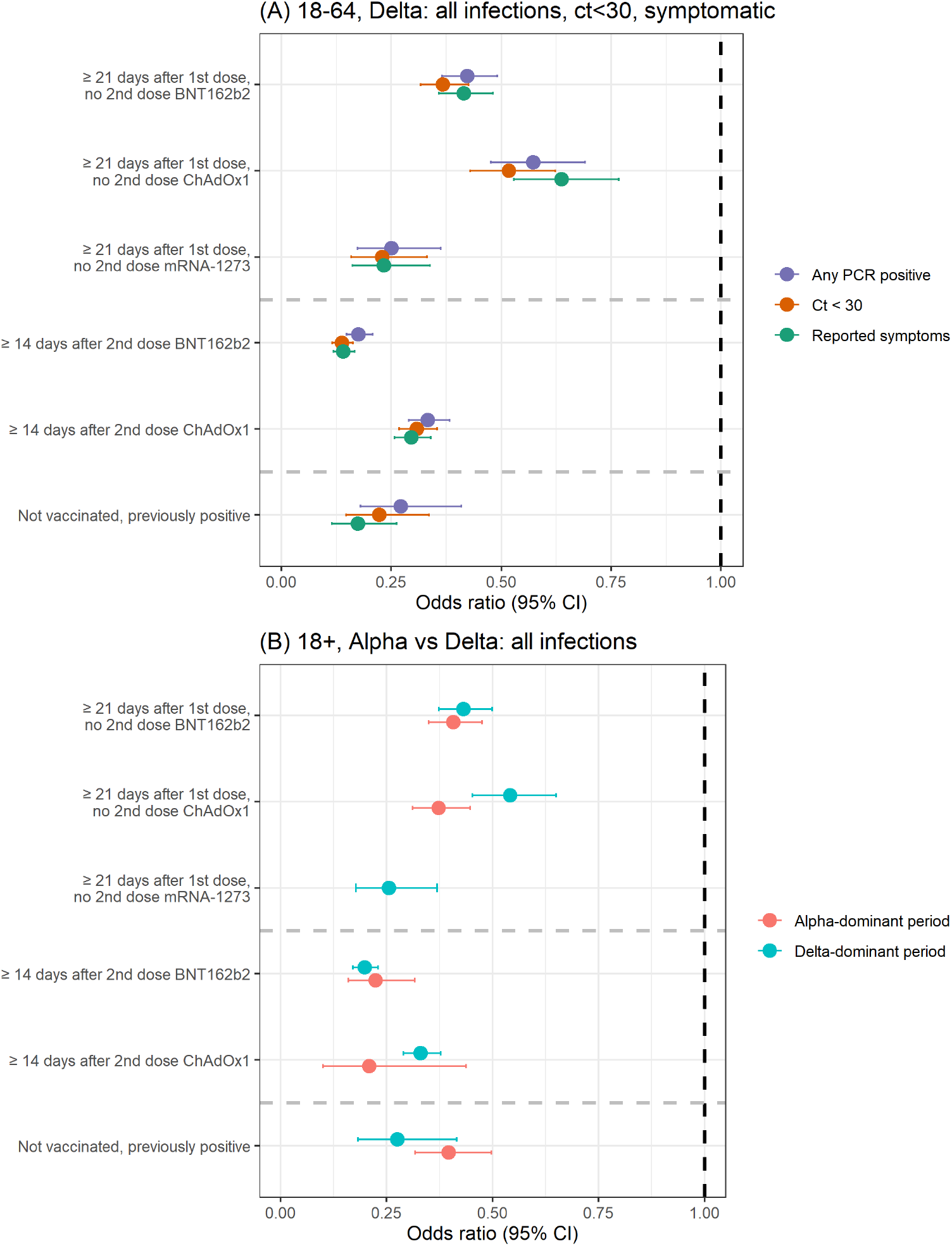
(A) Protection against all new PCR-positive episodes, those with Ct<30, or with self-reported symptoms in those 18-64 years in the Delta-dominant period (B) Protection against all new PCR-positive episodes in those 18+ years in both the Alpha- and Delta-dominant period. Note: Vaccine effectiveness estimates (=100%*(1-OR)) in **Table 1** and **2** for ≥18 years and 18 to 64 years respectively.

However, a decreasing number of visits remained in the unvaccinated reference group over time, particularly for older individuals (**Figure S2**). Whilst reasonable numbers of those aged 18 to 64 years remained in the unvaccinated reference group in the Delta-dominant period, comparisons with the Alpha-dominant period were not possible in this age group due to low numbers having received two vaccinations before 17 May 2021; however, VE estimates in the Delta-dominant period were similar to all adults for both vaccines (**Figure 1, Tables 1&2**). To investigate VE in the Delta-dominant period further, we therefore focussed on the younger age group.

In the Delta-dominant period, VE against new PCR-positives amongst those aged 18-64 years was significantly lower for ChAdOx1 versus BNT162b2 ≥21 days after one vaccination and ≥14 days after two vaccinations (heterogeneity p=0.001 and p<0.0001, respectively, **Table 2**). For both vaccines, having received two doses ≥14 days previously still provided significantly more protection compared with one dose ≥21 days previously (p<0.0001). There was no evidence that the effectiveness of two ChAdOx1 vaccinations ≥14 days previously in preventing new PCR-positives differed from the protection afforded by previous natural infection without vaccination (heterogeneity p=0.33), whereas two BNT162b2 vaccinations afforded greater protection (p=0.04). Results were similar for those ≥18y (**Table 1**).

**Table 2.**
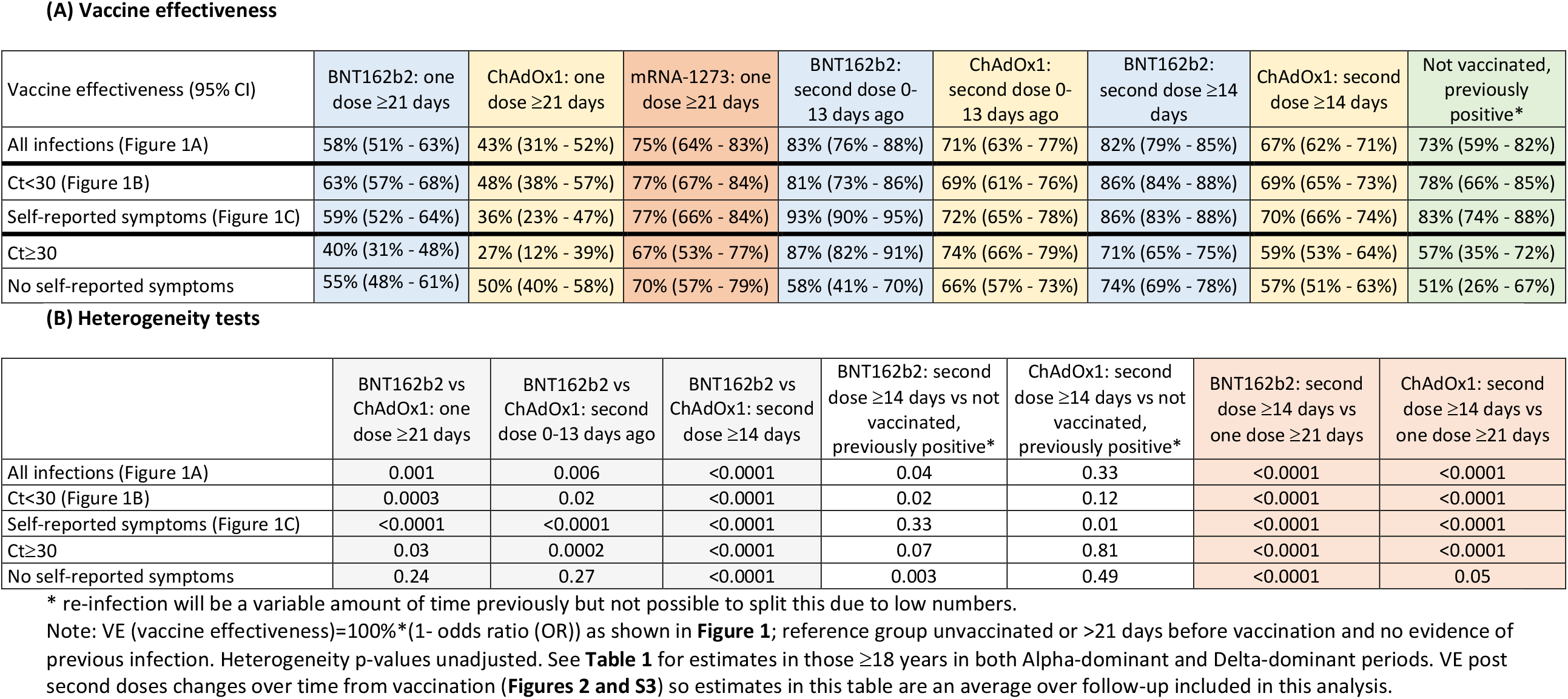
Vaccine effectiveness (VE) (95% CI) (A) and comparisons between vaccines and previous infection (B) for those 18 to 64 years in the Delta-dominant period.

| Vaccine effectiveness (95% CI) | BNT162b2: one dose ≥21 days | ChAdOx1: one dose ≥21 days | mRNA-1273: one dose ≥21 days | BNT162b2: second dose 0-13 days ago | ChAdOx1: second dose 0-13 days ago | BNT162b2: second dose ≥14 days | ChAdOx1: second dose ≥14 days | Not vaccinated, previously positive* |
| --- | --- | --- | --- | --- | --- | --- | --- | --- |
| All infections (Figure 1A) | 58% (51% - 63%) | 43% (31% - 52%) | 75% (64% - 83%) | 83% (76% - 88%) | 71% (63% - 77%) | 82% (79% - 85%) | 67% (62% - 71%) | 73% (59% - 82%) |
| Ct<30 (Figure 1B) | 63% (57% - 68%) | 48% (38% - 57%) | 77% (67% - 84%) | 81% (73% - 86%) | 69% (61% - 76%) | 86% (84% - 88%) | 69% (65% - 73%) | 78% (66% - 85%) |
| Self-reported symptoms (Figure 1C) | 59% (52% - 64%) | 36% (23% - 47%) | 77% (66% - 84%) | 93% (90% - 95%) | 72% (65% - 78%) | 86% (83% - 88%) | 70% (66% - 74%) | 83% (74% - 88%) |
| Ct≥30 | 40% (31% - 48%) | 27% (12% - 39%) | 67% (53% - 77%) | 87% (82% - 91%) | 74% (66% - 79%) | 71% (65% - 75%) | 59% (53% - 64%) | 57% (35% - 72%) |
| No self-reported symptoms | 55% (48% - 61%) | 50% (40% - 58%) | 70% (57% - 79%) | 58% (41% - 70%) | 66% (57% - 73%) | 74% (69% - 78%) | 57% (51% - 63%) | 51% (26% - 67%) |

**(B) Heterogeneity tests**
|  | BNT162b2 vs ChAdOx1: one dose ≥21 days | BNT162b2 vs ChAdOx1: second dose 0-13 days ago | BNT162b2 vs ChAdOx1: second dose ≥14 days | BNT162b2: second dose ≥14 days vs not vaccinated, previously positive* | ChAdOx1: second dose ≥14 days vs not vaccinated, previously positive* | BNT162b2: second dose ≥14 days vs one dose ≥21 days | ChAdOx1: second dose ≥14 days vs one dose ≥21 days |
| --- | --- | --- | --- | --- | --- | --- | --- |
| All infections (Figure 1A) | 0.001 | 0.006 | <0.0001 | 0.04 | 0.33 | <0.0001 | <0.0001 |
| Ct<30 (Figure 1B) | 0.0003 | 0.02 | <0.0001 | 0.02 | 0.12 | <0.0001 | <0.0001 |
| Self-reported symptoms (Figure 1C) | <0.0001 | <0.0001 | <0.0001 | 0.33 | 0.01 | <0.0001 | <0.0001 |
| Ct≥30 | 0.03 | 0.0002 | <0.0001 | 0.07 | 0.81 | <0.0001 | <0.0001 |
| No self-reported symptoms | 0.24 | 0.27 | <0.0001 | 0.003 | 0.49 | <0.0001 | 0.05 |

\* re-infection will be a variable amount of time previously but not possible to split this due to low numbers.
Note: VE (vaccine effectiveness)=100%\*(1- odds ratio (OR)) as shown in **Figure 1**; reference group unvaccinated or >21 days before vaccination and no evidence of previous infection. Heterogeneity p-values unadjusted. See **Table 1** for estimates in those ≥18 years in both Alpha-dominant and Delta-dominant periods. VE post second doses changes over time from vaccination (**Figures 2 and S3**) so estimates in this table are an average over follow-up included in this analysis.

### Delta-dominant period: impact of vaccination on new PCR-positives over time from second vaccination and in specific subgroups

In those 18 to 64 years, VE of BNT162b2 against new PCR-positives reduced by 22% (95% CI 6% to 41%) for every 30 days from second vaccination (p=0.007; **Figure 2**). Reductions were numerically smaller for ChAdOx1 (change -7% per 30 days, 95% CI -18% to +2%, p=0.15) but there was no formal evidence of heterogeneity (p=0.14).

**Figure 2.**
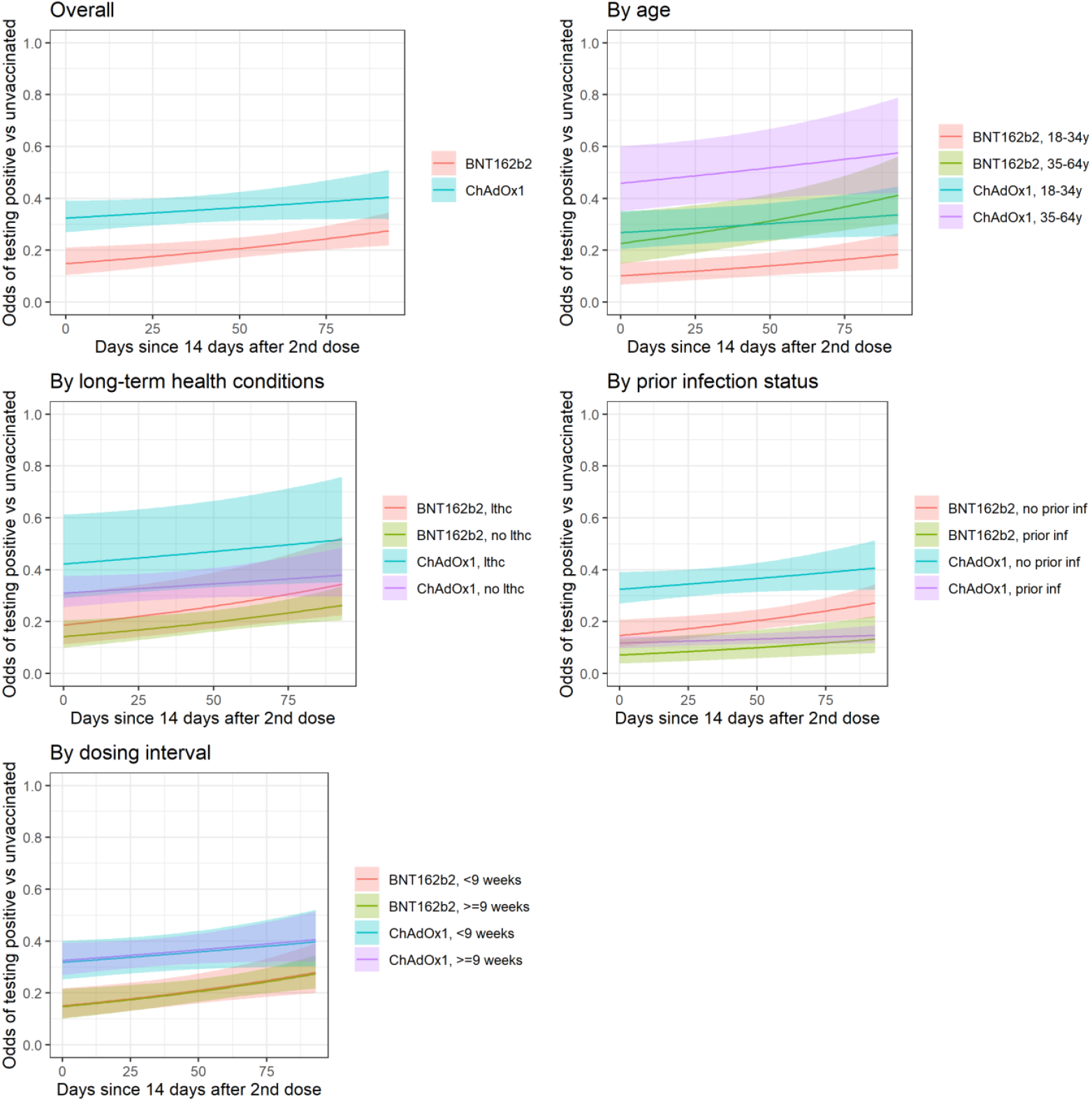
Protection against all new PCR-positive episodes over time from second dose, overall and by subgroups in those 18-64 years in the Delta-dominant period. Note: lthc=self-reporting a long term health condition. See **Figure S3** for effects on PCR-positive episodes with Ct<30. Overall odds ratio per 30 days longer from from ≥14 days after second vaccination 1.22 (95% CI 1.06-1.41) for BNT162b2 and for ChAdOx1 OR 1.07 (95% CI 0.98-1.18) (heterogeneity p=0.14). See **Table S3** for estimates of VE within subgroups 14 days after second vaccination (intercept on panels below).

Approximately 10% of visits in the Delta-dominant period occurred in vaccinated individuals with evidence of prior SARS-CoV-2 infection (**Table S2**). Protection against new PCR-positives was significantly higher for vaccinated individuals with prior infection than vaccinated individuals without prior infection (**Table S3**).

For example, 14 days after two ChAdOx1 vaccinations VE was 88% (95% CI 83-92%) among those with prior infection versus 68% (61-73%) in those without (heterogeneity p<0.0001); and 93% (87-96%) versus 85% (79-90%), respectively for BNT162b2 (heterogeneity p=0.006).

Vaccine effectiveness was also generally higher at younger ages (**Table S3**). For example, VE 14 days after the second BNT162b2 dose was 90% (85-93%) for those aged 18-34 years versus 77% (65-85%) for those aged 35-64 years (heterogeneity p=0.0001); and was 73% (65-80%) versus 54% (40-65%), respectively, for ChAdOx1 (heterogeneity p=0.002).

There was also no evidence of heterogeneity in VE between <9 versus ≥9 weeks between the first and second vaccination (approximate 25^th^ percentile) (heterogeneity p=0.80 and p=0.89 ≥14 days after two ChAdOx1 and BNT162b2 vaccinations, respectively, **Table S3)**. There was no evidence that the effect of vaccination on new PCR-positives differed between those reporting versus not reporting long-term health conditions (heterogeneity p>0.23 for BNT162b2 or Moderna, p>0.04 for ChAdOx1; **Table S3**).

### Impact of vaccination on new PCR-positives split by Ct and self-reported symptoms

Restricting new PCR-positives to those with Ct<30 (higher viral burden) or with symptoms, attenuations in VE in those ≥18 years in the Delta-dominant versus the Alpha-dominant period were more pronounced than against all new PCR-positives (**Table 1**). Importantly, attenuations in the Delta-dominant period now reached statistical significance for BNT162b2 as well as ChAdOx1 (e.g. Ct<30 VE ≥14 days post second dose 84% (82-86%) Delta versus 94% (91-96%) Alpha (heterogeneity p<0.0001), and 70% (65-73%) versus 86% (71-93%) respectively for ChAdOx1 (heterogeneity p=0.04)). In the Delta-dominant period, one or two BNT162b2 vaccinations still provided greater VE than ChAdOx1 against PCR-positives with Ct<30 or with reported symptoms in those ≥18 years (**Table 1**; p<0.003) and 18-64 years (**Figure 1, Table 2**; p<0.001). In the Delta-dominant period, VE against PCR-positives with Ct≥30 (lower viral burden) or without self-reported symptoms was only modestly lower than against PCR-positives with Ct<30 or with symptoms for all three vaccines (**Table 2**).

There was now formal evidence that the effectiveness of BNT162b2 against PCR-positives with Ct<30 or with symptoms declined faster ≥14 days after second vaccinations than for ChAdOx1 (heterogeneity p=0.003 for both outcomes; **Figure S3, Figure S4**). Extrapolating declines beyond the observed follow-up, both vaccines would be equally effective against PCR-positives with Ct<30 139 days (4.6 months) after the second dose and 116 days (3.8 months) against PCR-positives with symptoms.

### Viral burden and symptoms in new PCR-positives aged ≥18 years

In 12,287 new PCR-positives in the Alpha-dominant period, Ct values (inversely related to viral load) increased strongly with increasing time from first vaccination and number of doses (age/sex-adjusted trend-p<0.0001, **Figure 3A**). Ct values were highest in those ≥14 days after second vaccination (median (IQR) 33.3 (31.6-34.0) [N=56]); there was no evidence that this differed from those unvaccinated but previously PCR/antibody-positive (32.8 (30.9-34.2) [N=68]; age/sex-adjusted p=0.72), but Ct values were significantly higher than in those unvaccinated and not previously PCR/antibody-positive (28.7 (20.4-32.9) [N=10,853]; age/sex-adjusted p=0.02).

**Figure 3.**
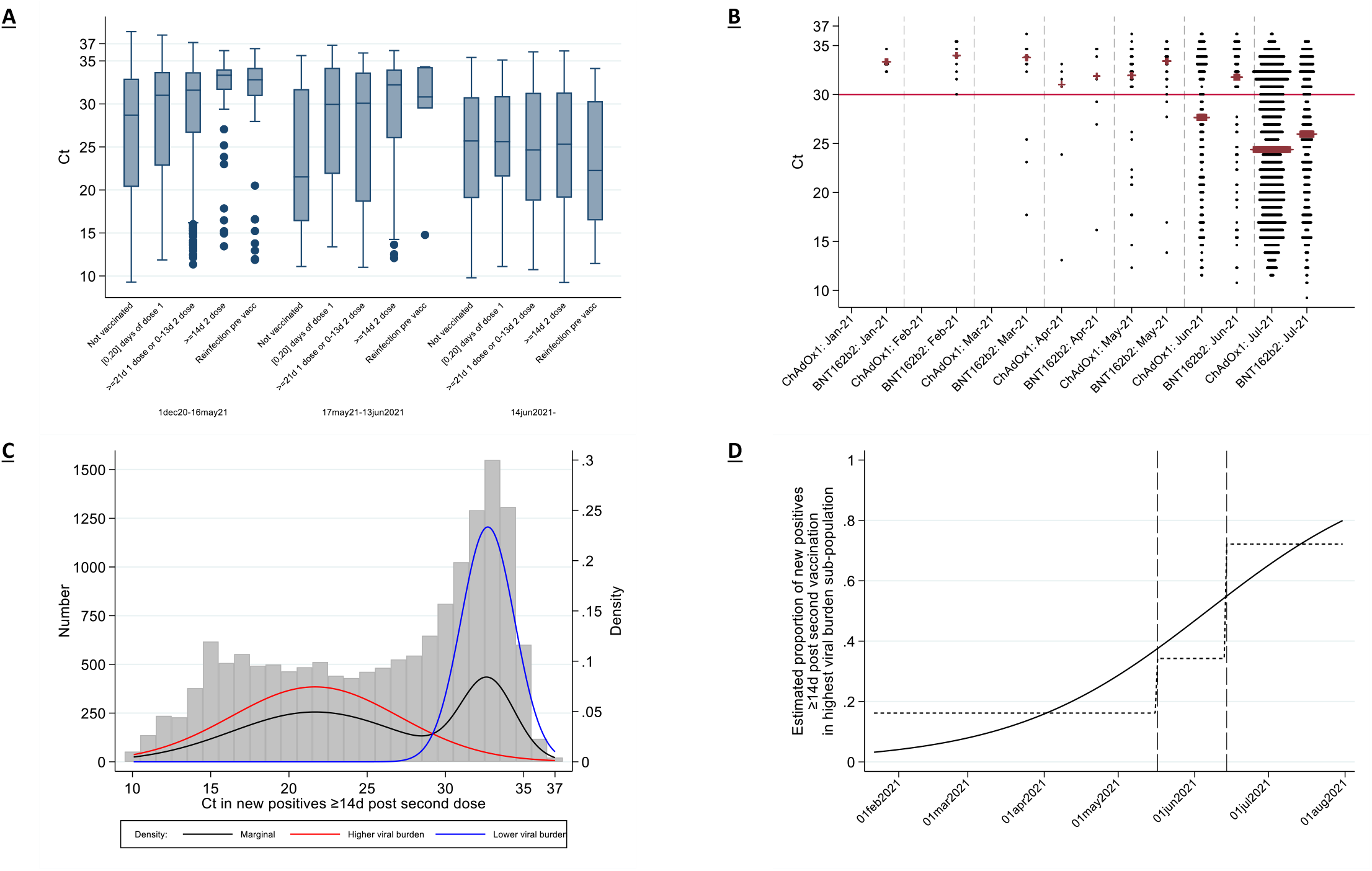
Ct values in new positive episodes in those 18y and older. (A) by vaccination/reinfection status and (B) vaccine type if ≥14 days after second dose over time. (C) shows observed Ct values from (B) with the marginal density (black) and the densities estimated from a two-component mixture distribution. (D) shows the probability that each new PCR-positive in (B) and (C) falls into the higher viral shedding class over time. Boxes in (A) are median (IQR).

From 14 June 2021, after which >92% of PCR-positives with Ct<30 were Delta-compatible (**Figure S1**), differences in Ct values between those unvaccinated (median (IQR) 25.7 (19.1-30.8) [N=326]) and ≥14 days after second vaccination (25.3 (19.1-31.3) [N=1593]) had attenuated substantially (age/sex-adjusted p=0.35, heterogeneity versus Alpha-dominant period p=0.01), as had differences with those unvaccinated but previously PCR/antibody-positive (22.3 (16.5-30.3) [N=20]). There was a trend towards lower Ct values in PCR-positives ≥14 days after two ChAdOx1 than two BNT162b vaccinations (**Figure 3B**; median -1.3 (95% CI -0.2 to +2.9), sex/age-adjusted p=0.08). Mirroring the attenuation in Ct values, the difference between those unvaccinated and ≥14 days after second vaccination in the percentages of PCR-positives reporting any or well-recognised COVID-19 symptoms (cough, fever, loss of taste/smell) significantly attenuated after 14 June 2021 (heterogeneity p<0.0001, p=0.008 respectively, **Figure S5**). However, this was likely driven by lower Ct values, as the association between Ct and symptom reporting remained broadly similar post-Delta (**Figure S6**).

Considering all 1,736 PCR-positives ≥14 days after two ChAdOx2 or BNT162b2 vaccinations (1,415 (82%) of which had ≥1 prior negative swabs after their second vaccination), Ct values came from a mixture of two sub-populations (**Figure 3C;** Bayesian Information Criterion 499.4 lower than single population). The low sub-population had mean Ct=21.7 (95%CI 21.2-22.2) and the high sub-population mean Ct=32.7 (32.5-33.0), consistent with either mild or late identified infection. The relative percentage of new PCR-positives falling into these two sub-populations varied strongly over time (p<0.0001; **Figure 3D**), with the percentage in the low Ct (high viral burden) sub-population averaging 16%, 34% and 72% through 16 May 2021, 17 May-13 June and 14 June onwards respectively.

Independently of this effect of calendar time (Alpha versus Delta), new PCR-positives were less likely to be in the low Ct sub-population 14 days after two BNT162b2 than ChAdOx1 vaccinations (adjusted odds ratio (aOR)=0.33 (95% CI 0.16-0.67) p=0.002; **Table S4, Figure S7A**), but this likelihood increased significantly over time from second vaccination (aOR per month=1.43 (1.07-1.91) p=0.01; unadjusted in **Figure 4A**). In contrast, there was no evidence of changing likelihood over time for ChAdOx1 (aOR per month=0.97 (0.79-1.19) p=0.78; heterogeneity p=0.02). Overall, therefore, by around 3 months post second vaccination the probability of being in the low Ct sub-population was similar for both BNT162b2 and ChAdOx1 (**Figure S7A**). Those previously PCR/antibody-positive were less likely to belong to the low Ct sub-population (p<0.0001), as were those reporting having long-term health conditions (p=0.006), potentially reflecting protection in the former and longer duration of PCR-positivity in the latter leading to late infections being more likely to be identified through the fixed testing schedule. There were no additional effects of sex, age (unadjusted in **Figure 4B**), or ethnicity on the probability of belonging to the low Ct sub-population (p>0.15).

**Figure 4.**
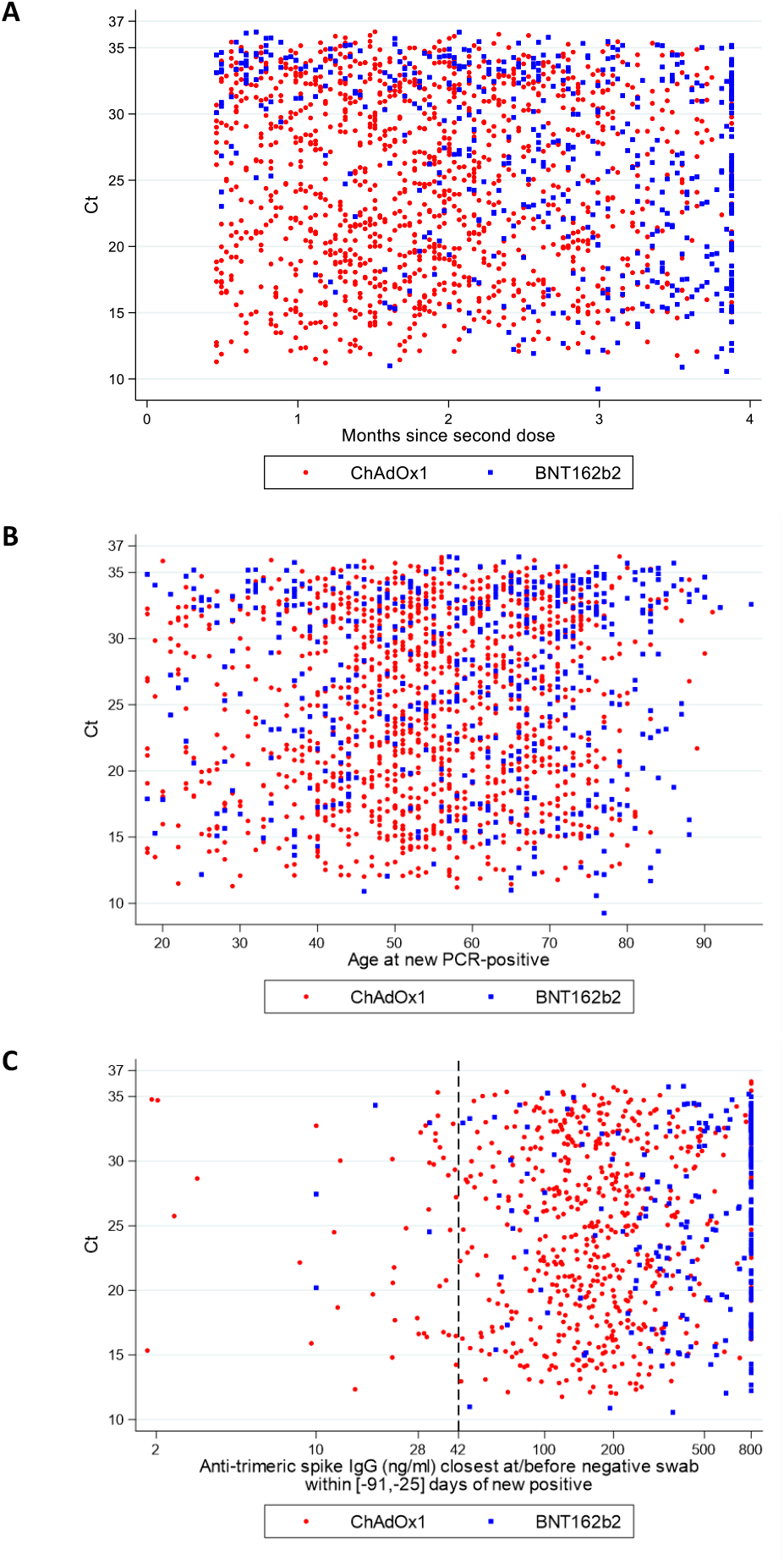
Ct values in new PCR-positives ≥14d after second ChAdOx1 or BNT162b2 vaccination by (A) months since second vaccination (N=1,736), (B) age (N=1,736) and (C) most recent anti-trimeric spike IgG antibody measurement (N=846). Note: antibody measurements taken median 30 (IQR 28-54) [range 25-91] days before the new PCR-positive, at or before the most recent prior negative swab and 14 days or more after first vaccination. 42ng/ml is the positivity threshold. Months since second dose truncated at 95^th^ percentile to avoid undue influence of outliers. Overall association with Ct Spearman rho=-0.09 (p=0.004) for time since second vaccination (A), 0.08 (p=0.002) for age (B), and 0.10 (p=0.002) for IgG (C).

Vaccine type and time from second vaccination had similar effects on the mean Ct within the low Ct sub-population, with higher Ct values in new PCR-positives 14 days after second BNT162b2 vaccination (p=0.003) which then dropped significantly faster with time from second vaccination than for ChAdOx1 (interaction p=0.01), leading to similar Ct values with both vaccines by around 3 months (**Figure S7B**). Calendar date was the only other factor strongly associated with Ct in both low and high Ct sub-populations (**Figure S7C**), with modest declines and increases within the low Ct sub-population consistent with increasing and decreasing positivity rates (**Figure S1C**) leading to new infections being identified slightly earlier/later^15^.

A prior anti-trimeric spike antibody (IgG) result was available for 846/1,736 (49%) new PCR-positives ≥14 days after two ChAdOx2 or BNT162b2 vaccinations, of which 795 (94%) were above the 42 ng/mL positivity threshold (**Figure 4C**), median 215 ng/ml (IQR 126-454). However, independently of factors in **Table S4**, every doubling in IgG was associated with 22% lower odds of a new PCR-positive belonging to the low Ct sub-population (aOR=0.78 (95% CI 0.66-0.93), p=0.007), with no evidence this varied by vaccine type (heterogeneity p=0.31). There was no evidence of association between IgG and mean Ct values within either sub-population (p>0.14). Most participants with antibody measurements after a new PCR-positive ≥14 days post second vaccination increased antibody levels after their new PCR-positive (**Figure S8**).

## Discussion

The results from this large community surveillance study show that vaccination with two doses of BNT162b2 or ChAdOx1 still significantly reduces the risk of new PCR-positive SARS-CoV-2 infections. However, whereas the two vaccines provided similar benefits when Alpha was dominant, benefits from two ChAdOx1 doses are reduced with Delta more than two BNT162b2 doses, although two ChAdOx1 doses still provide similar protection to that from previous natural infection. Benefits from both vaccines are numerically greater against PCR-positives with versus without self-reported symptoms and high versus low viral burden PCR-positives, but the difference in effectiveness is smaller with Delta for both vaccines.

The dynamics of protection varied over time from second vaccination, and by vaccine type, with initially larger effectiveness with BNT162b2 than ChAdOx1, which then become more similar by ∼4-5 months due to more rapid waning of effectiveness with BNT162b2, particularly against infections with Ct<30 or symptoms. Importantly, there was no evidence that effectiveness depended on the interval between first and second vaccinations (<9 weeks versus ≥9 weeks). Protection against new PCR-positives was significantly larger among those vaccinated with evidence of prior infection compared to those vaccinated without prior infection. While we could only assess the effectiveness of a single mRNA-1273 dose in the Delta-dominant period, this vaccine appeared more effective than a single dose of BNT162b2 or ChAdOx1, although this could potentially be driven by age, as those receiving mRNA-1273 were younger on average and effectiveness appeared greater in younger individuals. To date, other studies have only assessed mRNA-1273 effectiveness against other SARS-CoV-2 variants, and generally had limited power for comparisons with other vaccines or did not account for time since vaccination^1,17-21^. We also found greater effectiveness in those 18 to 34 than 35 to 64 years, although we were not able to jointly assess the degree to which this could have been cause by higher rates of previous infection in this group. We were unable to estimate vaccine effectiveness in those 65 years and older in the Delta-dominant period as very few individuals remained unvaccinated.

Few studies have assessed VE during periods where the Delta variant dominated. A test-negative case-control study from the English symptomatic testing program suggested that the effectiveness after one dose of either BNT162b2 or ChAdOx1 was lower against symptomatic infection with Delta than Alpha (31% versus 49%, respectively), with smaller differences after two doses (BNT162b2 88% versus 94%, respectively; ChAdOx1 67% versus 75%, respectively)^9^. There is little alternative to using observational data to assess vaccine effectiveness against new variants, since further placebo-controlled randomised trials would be unethical (although active comparator trials could still be performed). However, there are numerous biases in observational analyses^22^, particularly if symptomatic testing is non-random and related to perceived efficacy^10^. Potential bias due to such health-seeking behaviour is likely particularly pronounced for mild symptoms, included in many vaccine effectiveness studies using routine symptomatic testing program data. This may be exacerbated by the generic nature of many symptoms prompting testing, which may be incidental, and misclassification due to individuals reporting symptoms when they wish to get a test. As we demonstrated substantially lower VE against infections with high Ct or no reported symptoms, this would bias estimates towards lower effects, potentially differentially between vaccines.

Such bias is substantially reduced when testing schedules are fixed independent of symptom or vaccination status, as in our survey, or when using objective severe disease endpoints such as hospital admissions and deaths. A recent study from Scotland^11^ found no statistical evidence of differential effectiveness against hospital admissions with Delta and Alpha (62% versus 72% in PCR-positives), although power was relatively limited. BNT162b2 effectiveness against hospitalisations remained high when Delta dominated in Israel (88%)^23^, despite lower effectiveness against self-reported symptomatic SARS-CoV-2 infection (41% versus 97% previously)^24^.

Although testing behaviour bias could contribute to these differences, we also found a stronger protective effect against infections with higher viral burden and/or symptoms from BNT162b2 and ChAdOx1 vaccines, although to a lesser degree than against Alpha. One explanation could be differential effects of vaccination on mucosal and systemic immunity^25^. In theory, the former is more important for preventing carriage, transmission, and infection becoming established, while the latter is more important for preventing severe disease once infected^26^. Studies in rhesus macaques showed greater reductions in SARS-CoV-2 viral load in the lungs and prevention of pneumonia, without reducing viral loads in the upper respiratory tract with intramuscular ChAdOx1^27^, and protection against viral replication at much lower concentrations in the lower than the upper respiratory tract with intramuscular mRNA-1273^28^. In mice, an experimental adenovirus vaccine induced strong systemic adaptive immune responses against SARS-CoV-2 and reduced infection in the lungs, but minimal mucosal immune responses when administered intramuscularly^29^. Another explanation for differences in VE against infections with Delta versus Alpha is that the former may have a replication advantage in airway human epithelial cells; increased infectivity at mucosal surfaces could facilitate antibody evasion^30^. A final explanation could be varying protection by time since second vaccination in the Delta-dominant period, which also differed between BNT162b2 and ChAdOx1. When such time-dependent effects are present, studies with different follow-up will inevitably get different “average” results, and studies when Alpha dominated may predominantly reflect early effects. Regardless of explanation, whilst protection against hospitalisation and death is maintained, “booster” vaccinations may not be needed, particularly since infection post vaccination may provide a natural antibody boost. However, declines in immunity against infection demonstrate this needs to be monitored closely.

As well as reduced VE, we also found a substantial shift in viral burden in those infected despite two vaccinations with BNT162b2 or ChAdOx1 in the Delta-dominant period, with similar average Ct values to those infected without vaccination, and much more similar percentages reporting symptoms, driven by Ct. Whilst with Alpha, we^15^, and others^31-33^, found that vaccinated individuals had lower viral burden (higher Ct values) than unvaccinated individuals, the greater number of new PCR-positives (1,736 ≥14d post second vaccination) allowed us to show there are two different types of such infections, a low viral burden group that dominated early in 2021, and a high viral burden group which increased in frequency with Delta. Those receiving ChAdOx1 were more likely to fall into the latter group after their second vaccination, as were an increasing percentage of new PCR-positives with increasing time from second BNT162b2 vaccination, mirroring changes in protection against new PCR-positivity. Peak viral load therefore now appears similar in infected vaccinated and unvaccinated individuals, with potential implications for onward transmission risk, given the strong association between peak Ct and infectivity^34^. However, the degree to which this might translate into new infections is unclear; a greater percentage of virus may be non-viable in those vaccinated, and/or their viral loads may also decline faster as suggested by a recent study of patients hospitalised with Delta^31^ (supported by associations between higher Ct and higher antibody levels here and in^35^), leading to shorter periods “at risk” for onwards transmission. Nevertheless, there may be implications for any policies that assume a low risk of onward transmission from vaccinated individuals (e.g. relating to self-isolation, travel), despite vaccines both still protecting against infection, thereby still reducing transmission overall. This may be particularly important when vaccinated individuals are not aware of their infection status or perceive that their risk of transmission is low. Importantly, those infected after second vaccination appeared to gain an antibody boost, and higher prior antibody levels were independently associated with lower viral burden.

The main study strength is its size and design including participants from randomly selected private residential households in the community, tested following a fixed schedule, independent of symptoms and vaccination status, thereby avoiding bias due to test-seeking behaviour that potentially affects many other studies assessing vaccine effectiveness against SARS-CoV-2 infections^10^. Furthermore, we are able to adjust for risk factors that also affect vaccination but are typically not available in electronic health records, such as patient-facing healthcare work and long-term health-conditions, potentially leading to less residual confounding than studies relying on routine electronic healthcare data.

Our study has several limitations. While we have included a broad set of potential confounders, results may still be biased by unknown confounders or misclassification of prior infection status, for example due only having antibody measurements on a subset. Participants are tested initially at weekly and then monthly visits, meaning when rates are increasing, as when Delta came to dominate, we expect to identify infected individuals earlier in their infection episode^36,37^, as shown and adjusted for in our Ct analysis. Late detection of older infections on the fixed visit schedule means some positives could be classified as having occurred shortly after vaccination while the infection may actually have been acquired before vaccination, potentially diluting vaccine effectiveness estimates. However most infections ≥14d post second vaccination had a preceding negative after second vaccination. To avoid misclassification bias from erroneously classifying higher Ct positives where only ORF1ab+N genes were detected as Alpha, our comparisons treated calendar periods as an instrumental variable, according to whether Alpha or Delta was dominant, but this will likely lead to a small amount of bias in our vaccine effectiveness estimates.

In summary, with Delta, BNT162b2 and ChAdOx1 remain protective against any new PCR-positive and infections with higher viral burden or symptoms, but vaccine effectiveness is reduced, with evidence of significantly different dynamics of immunity against infections with Ct<30 or symptoms following second doses of the two vaccines. With Delta, those infections occurring despite either vaccine have similar peak viral burden to those in unvaccinated individuals. The impact on infectivity to others is unknown, but requires urgent investigation. It further argues for vaccinating as many of the population as possible, since those not vaccinated may not be protected by as substantial reductions in transmission among the immunised population as seen other infections, making herd immunity likely unachievable for emerging variants and requiring efforts to protect individuals themselves. Whilst the current preservation of VE against severe outcomes suggests that allowing ongoing virus transmission and nasopharyngeal viral presence may have limited consequences, the success of this strategy will ultimately rely on universal vaccination (currently not available to most worldwide), uniform protection induced by vaccines including in older individuals, optimisation of vaccine strategies to induce higher levels of mucosal and systemic immunity, and an absence of novel variants which might compromise VE against severe infection.

## Data Availability

Data are still being collected for the COVID-19 Infection Survey. De-identified study data are available for access by accredited researchers in the ONS Secure Research Service (SRS) for accredited research purposes under part 5, chapter 5 of the Digital Economy Act 2017. For further information about accreditation, contact or visit the SRS website.

https://www.ons.gov.uk/aboutus/whatwedo/statistics/requestingstatistics/approvedresearcherscheme

## Acknowledgements

This study is funded by the Department of Health and Social Care with in-kind support from the Welsh Government, the Department of Health on behalf of the Northern Ireland Government and the Scottish Government. EP, KBP, ASW, TEAP, NS, DE are supported by the National Institute for Health Research Health Protection Research Unit (NIHR HPRU) in Healthcare Associated Infections and Antimicrobial Resistance at the University of Oxford in partnership with Public Health England (PHE) (NIHR200915). ASW and TEAP are also supported by the NIHR Oxford Biomedical Research Centre. EP and KBP are also supported by the Huo Family Foundation. ASW is also supported by core support from the Medical Research Council UK to the MRC Clinical Trials Unit [MC_UU_12023/22] and is an NIHR Senior Investigator. PCM is funded by Wellcome (intermediate fellowship, grant ref 110110/Z/15/Z) and holds an NIHR Oxford BRC Senior Fellowship award. DWE is supported by a Robertson Fellowship and an NIHR Oxford BRC Senior Fellowship. The views expressed are those of the authors and not necessarily those of the National Health Service, NIHR, Department of Health, or PHE. The funder/sponsor did not have any role in the design and conduct of the study; collection, management, analysis, and interpretation of the data; preparation, review, or approval of the manuscript; and decision to submit the manuscript for publication. All authors had full access to all data analysis outputs (reports and tables) and take responsibility for their integrity and accuracy.

We are grateful for the support of all COVID-19 Infection Survey participants and the COVID-19 Infection Survey team:

Office for National Statistics: Sir Ian Diamond, Emma Rourke, Ruth Studley, Alex Lambert, Tina Thomas. Office for National Statistics COVID Infection Survey Analysis and Operations teams, in particular Daniel Ayoubkhani, Russell Black, Antonio Felton, Megan Crees, Joel Jones, Lina Lloyd, Esther Sutherland. University of Oxford, Nuffield Department of Medicine: Ann Sarah Walker, Derrick Crook, Philippa C Matthews, Tim Peto, Emma Pritchard, Nicole Stoesser, Karina-Doris Vihta, Alison Howarth, George Doherty, James Kavanagh, Kevin K Chau, Stephanie B Hatch, Daniel Ebner, Lucas Martins Ferreira, Thomas Christott, Brian D Marsden, Wanwisa Dejnirattisai, Juthathip Mongkolsapaya, Sarah Hoosdally, Richard Cornall, David I Stuart, Gavin Screaton.

University of Oxford, Nuffield Department of Population Health: Koen Pouwels.

University of Oxford, Big Data Institute: David W Eyre.

University of Oxford, Radcliffe Department of Medicine: John Bell.

Oxford University Hospitals NHS Foundation Trust: Stuart Cox, Kevin Paddon, Tim James.

University of Manchester: Thomas House. Public Health England: John Newton, Julie Robotham, Paul Birrell.

IQVIA: Helena Jordan, Tim Sheppard, Graham Athey, Dan Moody, Leigh Curry, Pamela Brereton. National Biocentre: Ian Jarvis, Anna Godsmark, George Morris, Bobby Mallick, Phil Eeles.

Glasgow Lighthouse Laboratory: Jodie Hay, Harper VanSteenhouse

## Author Contributions

The study was designed and planned by ASW, JF, JB, JN, ID and KBP and is being conducted by ASW, RS, DC and ER. This specific analysis was designed by ASW and KBP. KBP and ASW contributed to the statistical analysis of the survey data. JH conducted analysis of the RT-PCR data. ASW and KBP drafted the manuscript. All authors contributed to interpretation of the study results, and revised and approved the manuscript for intellectual content. KBP and ASW are the guarantors and accept full responsibility for the work and conduct of the study, had access to the data, and controlled the decision to publish. The corresponding author (KBP) attests that all listed authors meet authorship criteria and that no others meeting the criteria have been omitted.

## Methods

### Study participants

The Office for National Statistics (ONS) COVID-19 Infection Survey (CIS) is a large household survey with longitudinal follow-up (ISRCTN21086382, https://www.ndm.ox.ac.uk/covid-19/covid-19-infection-survey/protocol-and-information-sheets) (details in^14,15^). The study received ethical approval from the South Central Berkshire B Research Ethics Committee (20/SC/0195). Private households are randomly selected on a continuous basis from address lists and previous surveys to provide a representative sample across the UK. Following verbal agreement to participate, a study worker visited each selected household to take written informed consent for individuals aged 2 years and over. Parents or carers provided consent for those aged 2-15 years; those aged 10-15 years also provided written assent. For the current analysis we only included individuals aged 16 years and over who were potentially eligible for vaccination.

Individuals were asked about demographics, behaviours, work, and vaccination uptake (https://www.ndm.ox.ac.uk/covid-19/covid-19-infection-survey/case-record-forms). At the first visit, participants were asked for (optional) consent for follow-up visits every week for the next month, then monthly for 12 months from enrolment. At each visit, enrolled household members provided a nose and throat self-swab following instructions from the study worker. From a random 10-20% of households, those 16 years or older were invited to provide blood monthly for antibody testing from enrolment. From April 2021, additional participants were invited to provide blood samples monthly to assess vaccine responses, based on a combination of random selection and prioritisation of those in the study for the longest period (independent of test results). Throughout, participants with a positive swab test and their household members were also invited to provide blood monthly for follow-up visits after this.

### Laboratory testing

Swabs were couriered directly to the UK’s national Lighthouse laboratories (Glasgow and the National Biocentre in Milton Keynes (to 8 February 2021)) where samples were tested within the national testing programme using identical methodology. The presence of three SARS-CoV-2 genes (ORF1ab, nucleocapsid protein (N), and spike protein (S)) was identified using real-time polymerase chain reaction (RT-PCR) with the TaqPath RT-PCR COVID-19 kit (Thermo Fisher Scientific, Waltham, MA, USA), analysed using UgenTec Fast Finder 3.300.5 (TagMan 2019-nCoV assay kit V2 UK NHS ABI 7500 v2.1; UgenTec, Hasselt, Belgium). The assay plugin contains an assay-specific algorithm and decision mechanism that allows conversion of the qualitative amplification assay raw data into test results with little manual intervention. Samples are called positive if either N or ORF1ab, or both, are detected. The S gene alone is not considered a reliable positive, but could accompany other genes (ie, one, two, or three gene positives).

Blood samples were couriered directly to the University of Oxford, where they were tested for the SARS-CoV-2 antibody using an ELISA detecting anti-trimeric spike IgG^38^. Before 26 February 2021, the assay used fluorescence detection as previously described (positivity threshold 8 million units). After this, it used a commercialised CE-marked version of the assay, the Thermo Fisher OmniPATH 384 Combi SARS-CoV-2 IgG ELISA (Thermo Fisher Scientific, Waltham, MA, USA), with the same antigen and a colorimetric detection system (positivity threshold 42 ng/ml monoclonal antibody unit equivalents, determined from 3840 samples run in parallel). From 27 February 2021, samples were also tested using a Thermo Fisher N antibody.

### Inclusion and exclusion criteria

This analysis included participants aged 18 years or over (i.e. those who were eligible for vaccination), and all visits with positive or negative swab results from 1 December 2020 to 1 August 2021. The analysis of vaccine effectiveness comparing Alpha-dominant and Delta-dominant periods included all individuals ≥18 years; analyses of the Delta-dominant period were also restricted to visits in those aged 18 to 64 years, as the vast majority of those 65 years and older were vaccinated twice before Delta became dominant (**Figure S2**). Analyses of Ct values in new PCR-positives by vaccination status included all individuals ≥18 years.

### Vaccination status

Participants were asked about their vaccination status at visits, including type, number of doses and date(s). Participants from England were also linked to administrative records from the National Immunisation Management Service (NIMS). We used records from NIMS where available, otherwise records from the survey, since linkage was periodic and NIMS does not contain information about vaccinations received abroad or in Northern Ireland, Scotland, and Wales. Where records were available in both, agreement on type was 98% and on dates 95% within ±7 days. A small number of visits after reported vaccination with either unknown or vaccines other than ChAdOx1, BNT162b2 or mRNA-1273 (for the latter we only included the first dose and only for the period ≥17 May) were excluded as these were too few to provide reliable estimates.

### SARS-CoV-2 positive episodes

PCR-positive results may be obtained at multiple visits after infection, so we grouped positive tests into ‘episodes’. Whole genome sequencing is available on only a subset of positives, and only a subsample provide monthly blood samples for antibody status, so positive episodes were defined using study PCR results. We previously found that defining episodes based on 90 days as suggested by the World Health Organisation (WHO)^39^ led to higher than plausible risk of a new episode between 90-120 days, particularly for high Ct infections^15^, suggesting intermittent long-term PCR positivity could be contributing. Here, we therefore defined the start of a new ‘positive episode’ as the date of either: i) the first PCR-positive test in the study (not preceded by any study PCR-positive test by definition); ii) a PCR-positive test after 4 or more consecutive negative tests; or iii) a PCR-positive test at least 120 days after the start of a previous episode with one or more negative tests immediately preceding this. Positive episodes were used to classify exposure groups and outcomes (see below).

### Exposures

At each study visit, a participant was classified into one of 13 different exposure groups based on current vaccination status, study antibody and PCR tests, and (for exposure classification only) positive swab tests linked from the English national testing programme^40^ (prior to visit), as follows:

i. Visits from participants ≥21 days before first vaccination, including those currently with no vaccination date, with no prior PCR or antibody-positive in the study, nor a positive swab test in the national testing programme (as defined below) (“Not vaccinated, not previously positive, ≥21 days before vaccination”) (baseline group);
ii. Visits from participants 1 to 21 days before first vaccination with no prior PCR or antibody-positive in the study, nor a positive swab test in the national testing programme (“Not vaccinated, not previously positive, 1-21 days before vaccination”)
iii. Visits 0 to 20 days following a first vaccination with BNT162b2 (“Vaccinated 0-20 days ago BNT162b2”);
iv. Visits 0 to 20 days following a first vaccination with ChAdOx1 (“Vaccinated 0-20 days ago ChAdOx1”);
v. Visits 0 to 20 days following a first vaccination with mRNA-1273 (“Vaccinated 0-20 days ago mRNA-1273”);
vi. Visits 21 days or more following a first vaccination with BNT162b2 but before a second vaccination (“≥21 days after 1^st^ dose, no second vaccination BNT162b2”);
vii. Visits 21 days or more following a first vaccination with ChAdOx1 but before a second vaccination (“≥21 days after 1^st^ dose, no second vaccination ChAdOx1”);
viii. Visits 21 days or more following a first vaccination with mRNA-1273 but before a second vaccination (“≥21 days after 1^st^ dose, no second vaccination mRNA-1273”);
ix. Visits 0 to 13 days following a second vaccination with BNT162b2 (“2^nd^ dose 0-13 days ago BNT162b2”);
x. Visits 0 to 13 days following a second vaccination with ChAdOx1 (“2^nd^ dose 0-13 days ago ChAdOx1”);
xi. Visits ≥14 days following second vaccination with BNT162b2 (“≥14 days after 2^nd^ dose BNT162b2”);
xii. Visits ≥14 days following second vaccination with ChAdOx1 (“≥14 days after 2^nd^ dose ChAdOx1”);
xiii. Visits from participants that had not yet been vaccinated but were previously PCR/antibody positive in the study, or had a positive swab test in the national testing programme based on the definition of positive episodes above (“Not vaccinated, previously positive”).

We chose these vaccination status categories empirically based on previous findings^15^. Exposure group ii (Not vaccinated, not previously positive, 1-21 days before vaccination) was included because there is inevitably a degree of transient reverse causality where vaccination appointments have to be rescheduled if someone tests positive in the weeks before the scheduled visit. As antibody status before vaccination is not available for all participants, we defined prior positivity by having either a previous PCR-positive episode or a positive S-antibody measurement >90 days before the visit or two consecutive positive N-antibody measurements >42 days before the visit. The choice of 90 and 42 days was arbitrary, but designed to exclude ongoing infections acquired previously being misattributed to current visits. Visits from vaccinated individuals (groups (iii)-(xii)) were defined irrespective of previous positivity (**Table S2**) to reflect the impact of vaccination as being implemented in the UK (without regard to prior infection). However, in sensitivity analysis we analysed the impact of vaccination by prior infection status. Visits from the same participant were classified in different groups depending on their status at each visit.

### Outcomes

Analysis was based on visits, since these occur independently of symptoms and are therefore unbiased. Only the first test-positive visit in each new PCR-positive infection episode starting after 1 December 2020 was used, dropping all subsequent visits in the same infection episode and all negative visits before the first time a participant could be considered “at risk” for a subsequent new positive episode (as defined above), to avoid misattributing ongoing PCR-positivity to visit characteristics and immortal time bias respectively. Primary analysis included all new PCR-positive episodes. Secondary analyses considered infection severity, by classifying positives by cycle threshold (Ct) value (<30 or ≥30) and self-reported symptoms. The threshold Ct value of 30 is somewhat arbitrary, but corresponds to ∼150 copies/ml^34^, and is consistently used in the UK for many purposes, including algorithms for review of low level positives at the laboratories where the PCR tests were performed and a threshold for attempting whole genome sequencing. For each positive test, a single Ct was calculated as the arithmetic mean across detected genes (Spearman correlation>0.98), then the minimum value was taken across positives in the infection episode to reflect the greatest measured viral burden within an episode. To allow for pre-symptomatic positives being identified in the survey, any self-reported symptoms at any visit within 0 to 35 days after the index positive in each infection episode were included (questions elicit symptoms in the last 7 days at each visit). Finally, positive infection episodes were classified as triple positive (ORF1ab+N+S or ORF1ab+S or N+S at least once across the episode; Delta-compatible), positive only for ORF1ab+N across the episode and never S-positive (Alpha-compatible, since Alpha has deletions in the S gene leading to S gene target failure) or always positive only on a single gene. As S-gene target failure may also occur in high Ct samples, the main analysis considered two periods of time when Alpha dominated (1 December 2020 to 16 May 2021) and when Delta dominated (17 May 2021 onwards) (**Figure S1**), further dividing analysis of Ct values at 14 June 2021.

### Confounders

The following potential confounders were adjusted for in all models as potential risk factors for acquiring SARS-CoV-2 infection: geographic area and age in years (see below), sex, ethnicity (white versus non-white as small numbers), index of multiple deprivation (percentile, calculated separately for each country in the UK)^41-44^, working in a care-home, having a patient-facing role in health or social care, presence of long-term health conditions, household size, multigenerational household, rural-urban classification^45-47^, direct or indirect contact with a hospital or care-home, smoking status, and visit frequency. Details are shown in **Table S1**.

### Statistical analysis

Associations between the different exposure groups and outcome (first positive test in an infection episode versus test-negative) were evaluated with generalised linear models with a logit link. Robust standard errors were used to account for multiple visits per-participant. To adjust for substantial confounding by calendar time and age, with non-linear effects of age which are also different by region, we included both as restricted cubic splines and interactions between these splines and region/country (regions for England and country for Northern Ireland, Scotland and Wales). Furthermore, given previous observations of different positivity rates by age over time^14^, we added a tensor spline to model the interaction between age and calendar time with the restriction that the interaction is not doubly non-linear^48^. The primary analysis considered effect modification of each vaccine exposure group by time period (before (Alpha-dominant) or after (Delta-dominant) 17 May 2021) in those aged ≥18 years. Secondary analyses considered variation over time from second vaccination (linear on the log-odds scale, truncating at the 95^th^ percentile of observed days from second vaccination separately for each vaccine), and effect modification by long-term health conditions, dosing interval, and prior infection status in the Delta-dominant period only in those aged 18 to 64 years. Pairwise comparisons of the exposure groups were performed unadjusted. Analysis was based on complete cases (>99% observations). Comparisons of Ct values by vaccine exposure groups used quantile (median) regression adjusted for age and sex. Associations between factors and Ct values were assessed using mixture normal linear regression models. We conducted backwards elimination (exit p=0.05) for associations between factors and the latent class probabilities and separately with the Ct values in each sub-population for the 12 variables shown in **Table S4**. We included interactions with vaccine in either part of the model type where these had interaction p<0.05. We considered three knot restricted natural cubic splines in contiuous factors (calendar date of positive, age, interval between first and second vaccination, time since second vaccination) (knots at the 10^th^, 50^th^ and 95^th^ percentiles) if there was evidence of non-linearity at p<0.01. To reduce the influence of outliers, we truncated the interval between first and second vaccination at 3 and 14 weeks, and the time from second vaccination at the 95^th^ percentile (118 days, 3.9 months).

## Code Availability

All statistical analyses of vaccine effectiveness were performed using standard functions in the following R packages: ggplot2 (version 3.3.2), rms (version 6.0-1), dplyr (version 1.0.2), emmeans (version 1.5.1), haven (version 2.3.1), sandwich (version 3.0-0), ggeffects (version 1.0.1), broom (version 0.7.2), multcomp (version 1.4-14), and Epi (version 2.44)). Analyses of Ct values were performed using qreg and fmm in Stata v16.1.Code used for data analysis is available upon request.

## Supplementary Material

**Figure S1.**
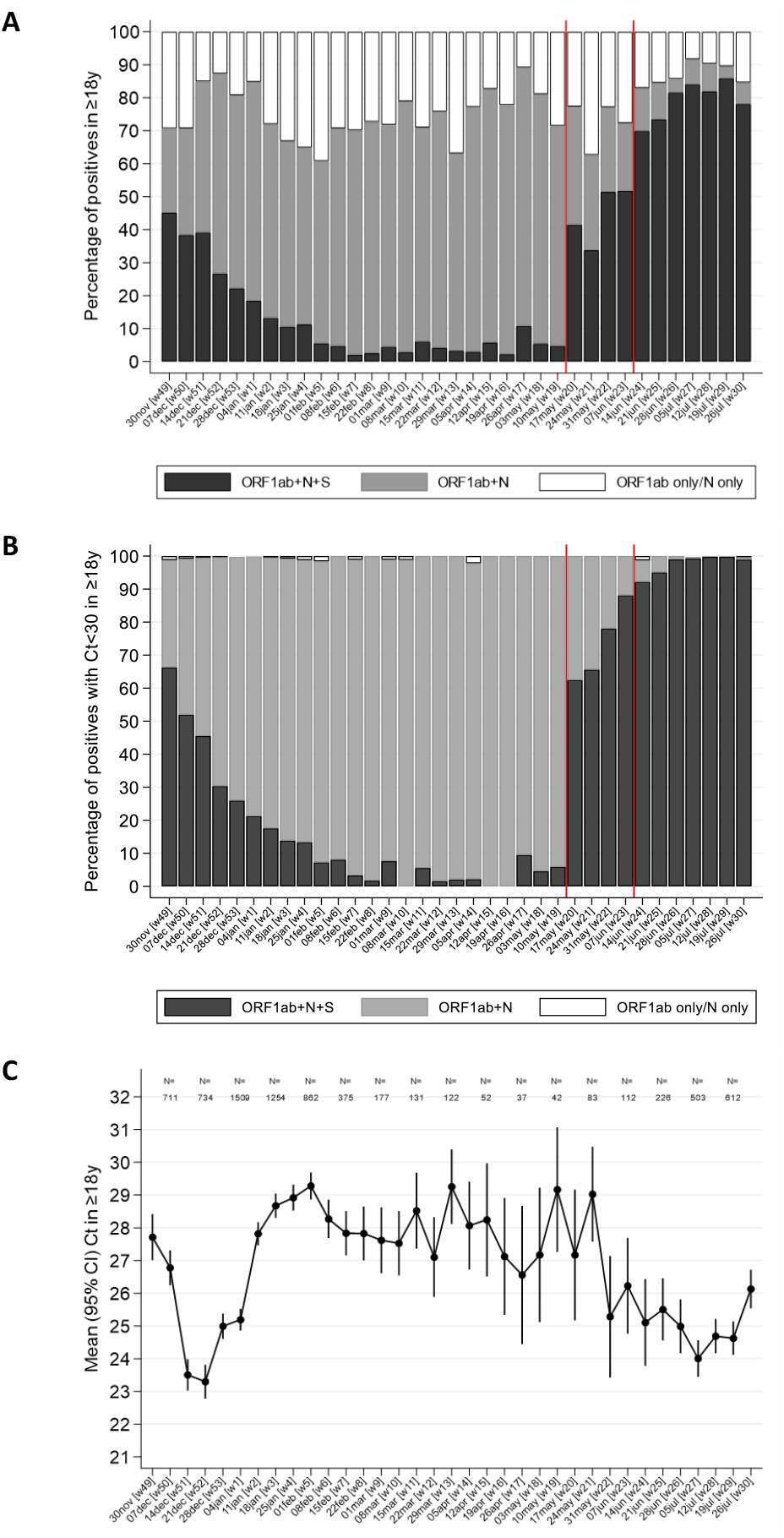
In new PCR-positive episodes in those ≥18 years, gene positivity pattern overall (A) and restricted to episodes with cycle threshold (Ct) <30 (B), and mean Ct value in all positives (C). Note: ORF1ab+N+S (black) are compatible with wild-type and Delta variants (S-gene positive); ORF1ab+N (gray) are compatible with the Alpha variant (S-gene negative). Those PCR-positives where only a single gene (N or ORF1ab were detected) cannot be classified (vast majority Ct>30). The percentage of PCR-positives with Ct<30 that were ORF1ab+N+S, compatible with Delta, increased from 6% the week commencing 10 May 2021, to 67% and 92% the weeks starting 17 May and 14 June 2021, respectively.

**Figure S2.**
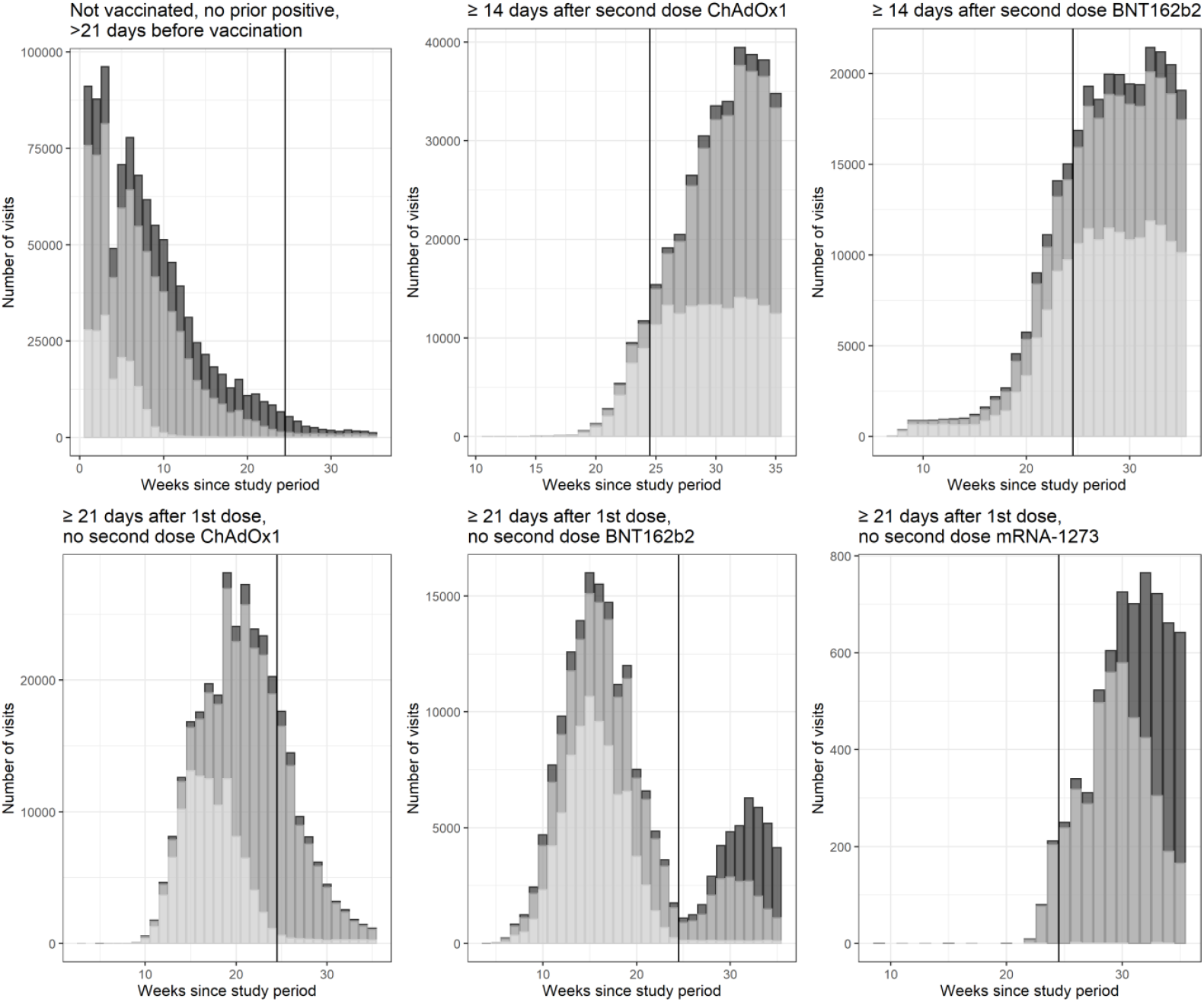
Visits included in analysis over time by vaccination status. Note: The graphs (with different scales for the axes) show the number of visits by vaccination status, by calendar time and age category (dark: 18-34 year olds, intermediate: 35-64 year olds; light: 65+ year olds. The vertical line at 25 weeks indicates the start of the period dominated by Delta.

**Figure S3.**
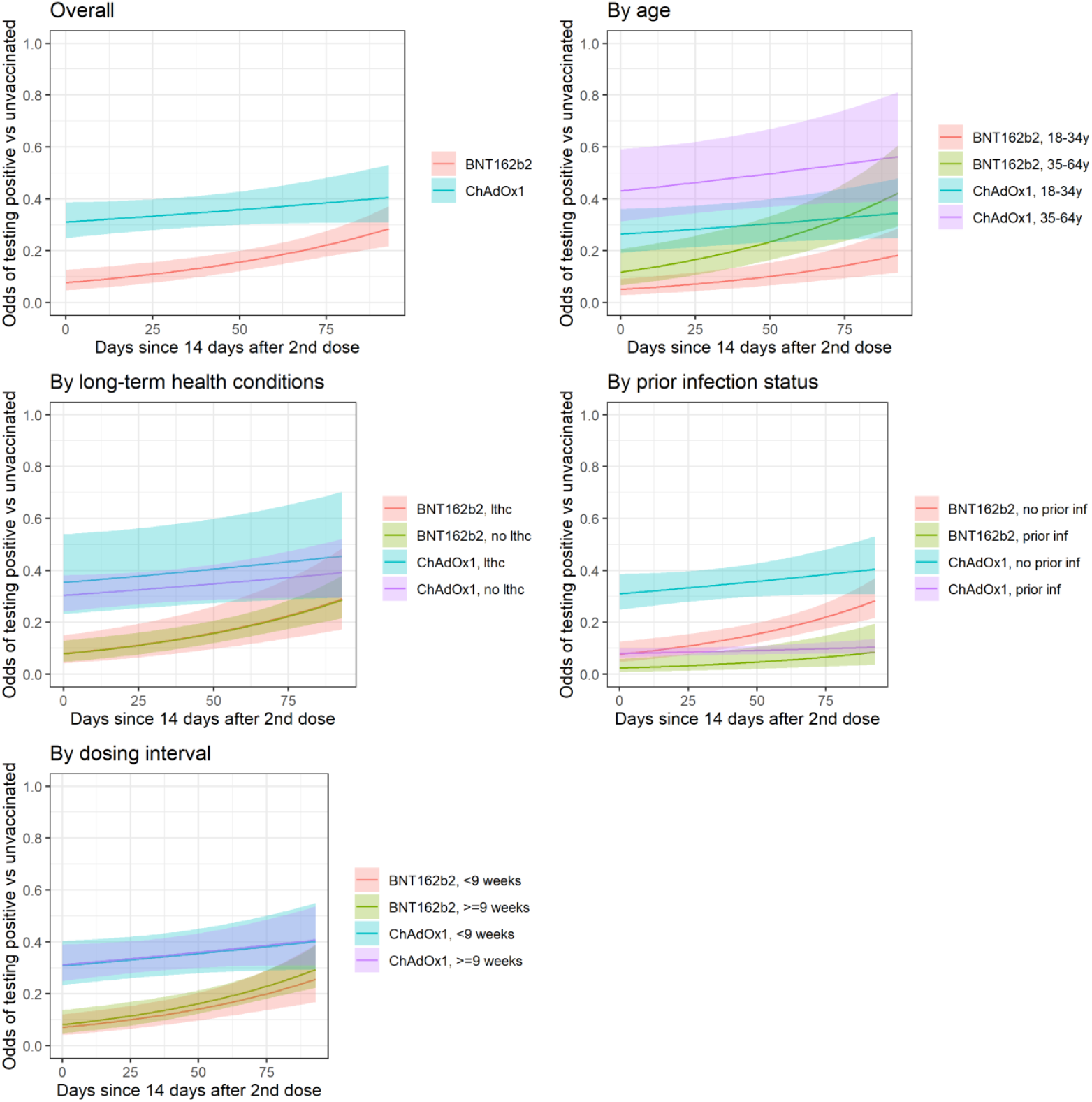
Protection against all new PCR-positive episodes with Ct<30 over time from second dose, overall and by potential subgroups in those 18-64 years in the Delta-dominant period. Note: lthc=self-reporting a long term health condition. See **Figure 2** for effects on all PCR-positive episodes. Overall odds ratio per 30 days longer from ≥14 days after second vaccination 1.52 (95% CI 1.26 to 1.84) (p<0.0001) for BNT162b2 and for ChAdOx1 OR=1.09 (95% CI 0.0.97 to 1.22) (p=0.14) (heterogeneity p=0.003). See **Table S3** for estimates of VE within subgroups 14 days after second vaccination (intercept on panels below).

**Figure S4.**
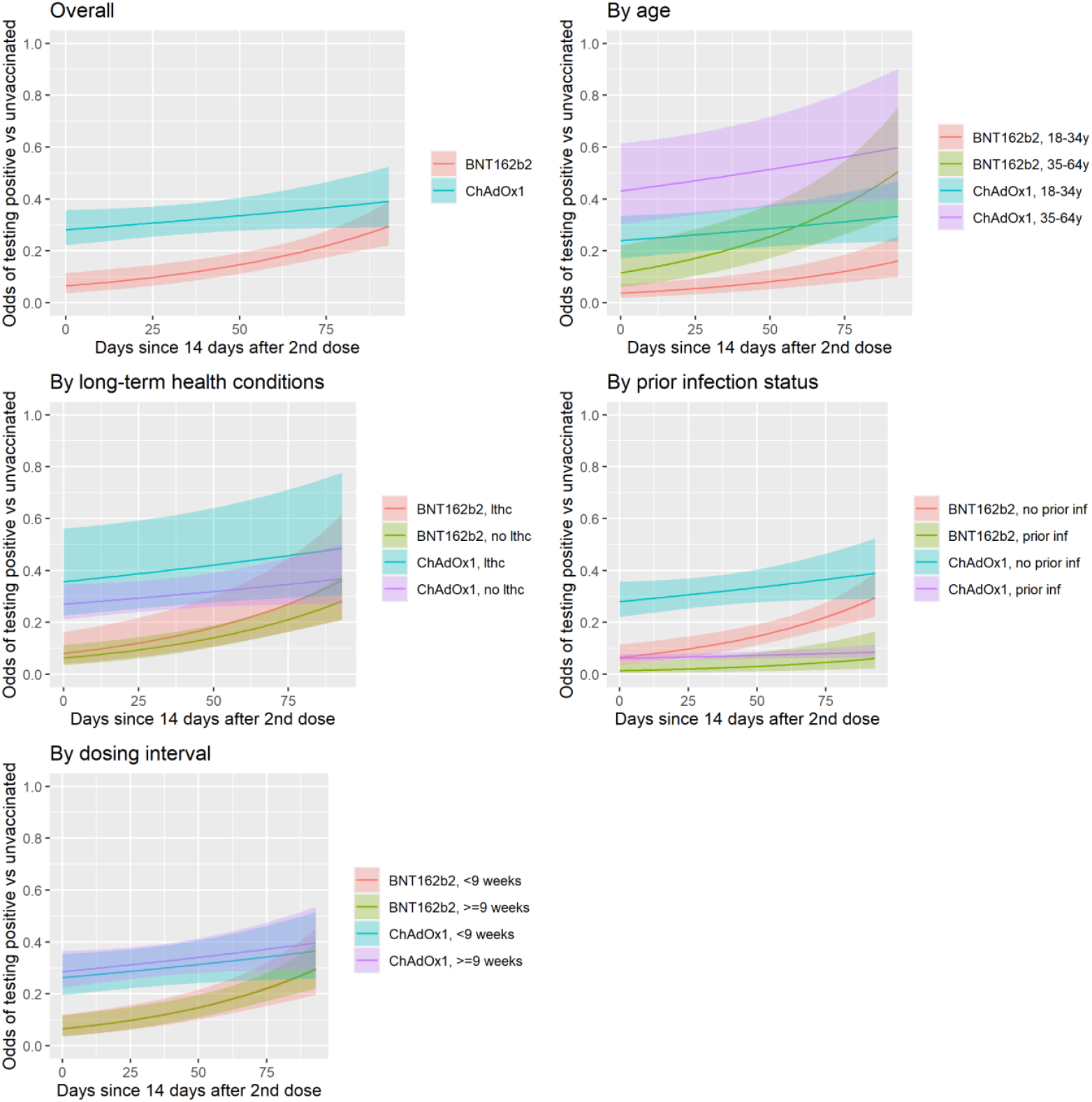
Protection against all new PCR-positive episodes with reported symptoms over time from second dose, overall and by potential subgroups in those 18-64 years in the Delta-dominant period. Note: lthc=self-reporting a long term health condition. See **Figure 2** for effects on all PCR-positive episodes. See **Table S3** for estimates of VE within subgroups 14 days after second vaccination (intercept on panels below).

**Figure S5.**
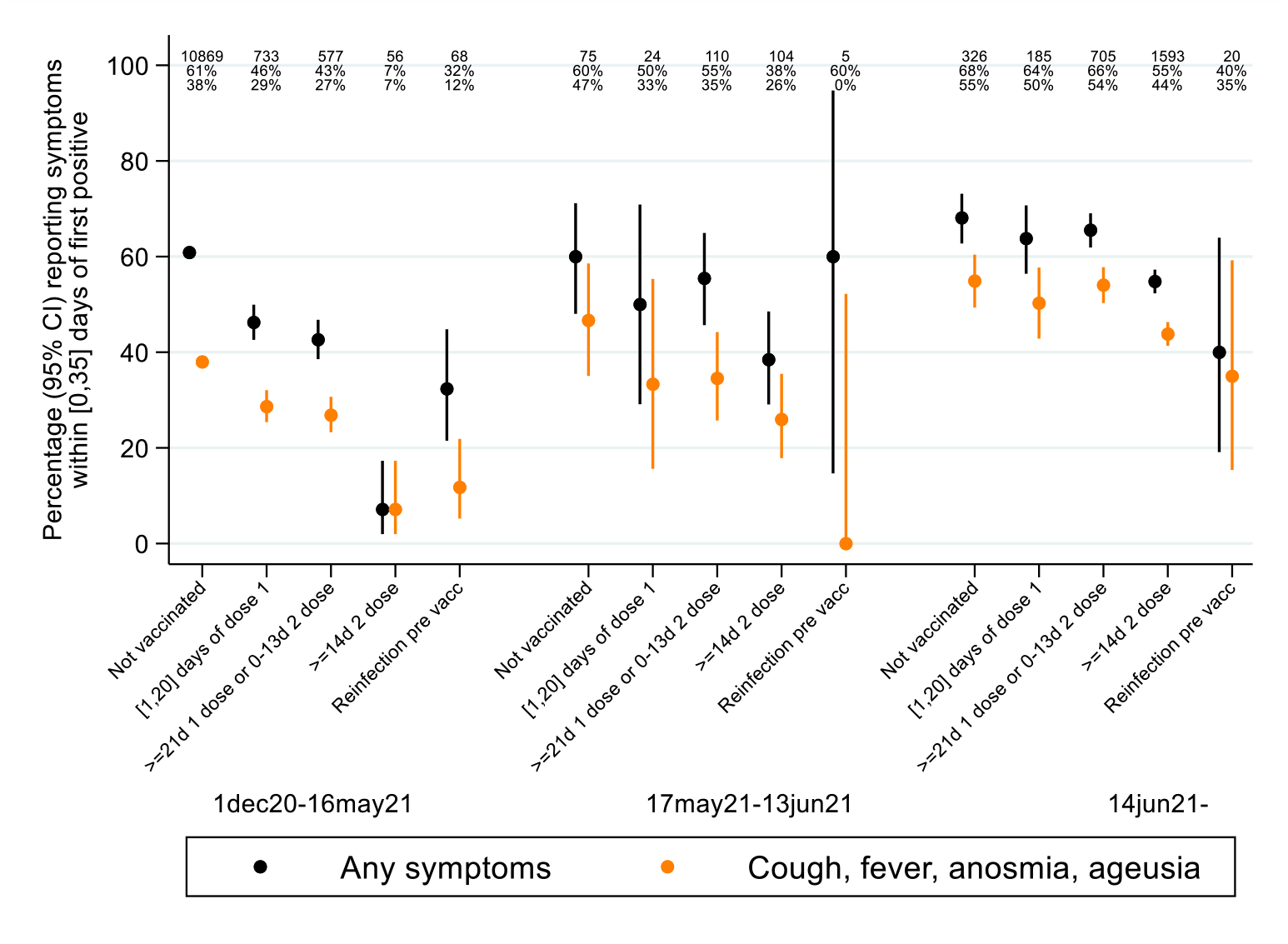
Symptoms reported in new PCR-positives aged ≥18 years by vaccination/reinfection status.

**Figure S6.**
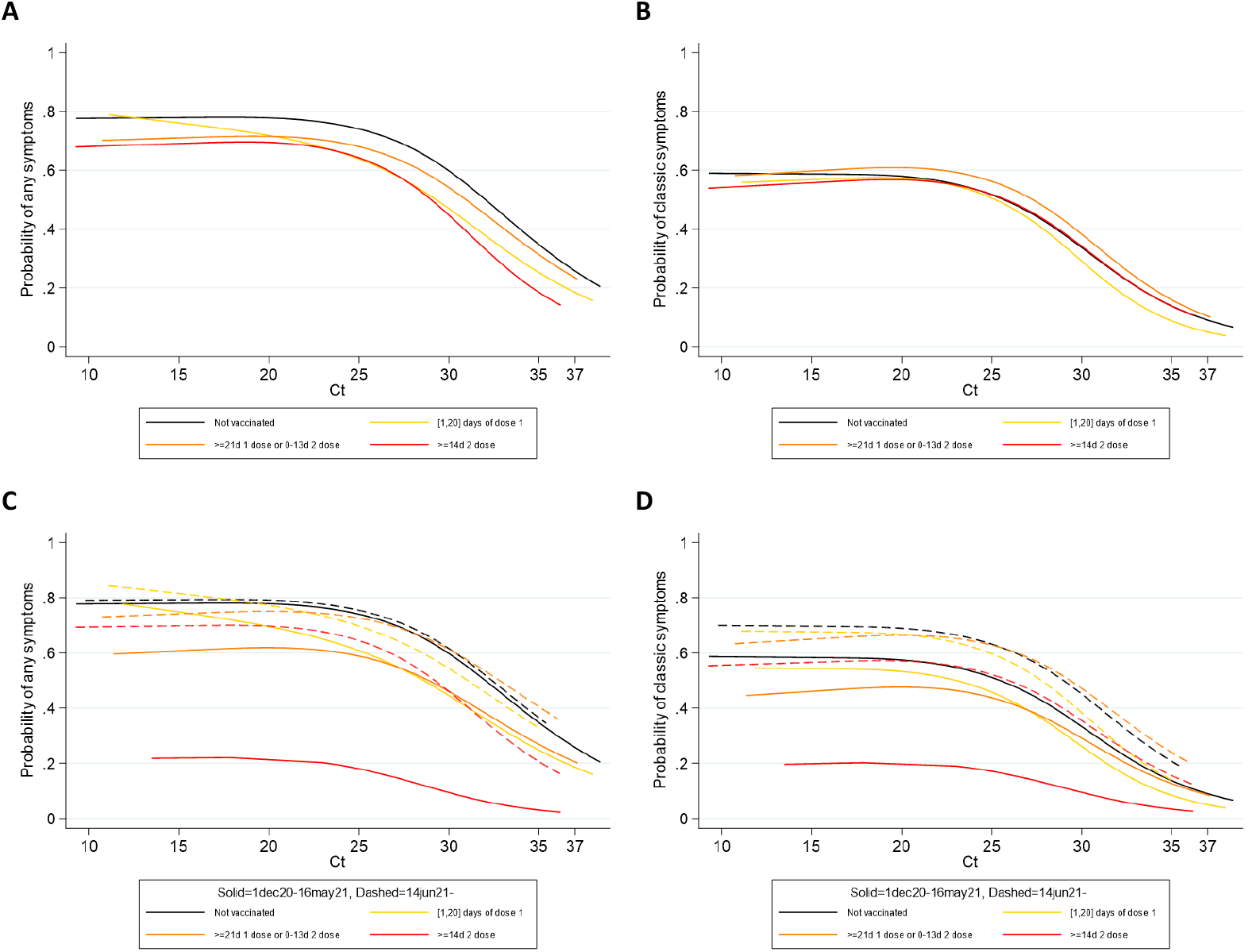
Probability of reporting any (A, C) or classic (cough, fever, loss of taste/smell) (B, D) symptoms by Ct value and vaccination status in new PCR-positives aged ≥18 years. Panels (A) and (B) include all PCR-positives from 1 December 2020 to 1 August 2021; panels (C) and (D) fit separate models to the periods 1 December 2020-16 May 2021 (solid lines) and 14 June 2021-1 August 2021 (dashed lines). Over the whole study period there was no strong evidence that the association between Ct values and probability of reporting symptoms differed across vaccine groups (any symptoms: heterogeneity p=0.04 (all models also adjusting for sex and age as a 5-knot restricted natural cubic spline), classic: p=0.58) although the absolute probability of reporting symptoms at all was slightly lower in those post-vaccination for any symptoms (p=0.007) but not classic symptoms (classic p=0.48). This could potentially reflect reporting bias. There were small numbers of new PCR-positives ≥14d post second vaccination before 17 May 2021, and symptom reporting was very low (red line, panels C and D), again potentially reflecting reporting bias given perceptions of vaccine efficacy. Comparing the periods before 17 May and after 14 June 2021, those ≥21 days post first vaccination but <14d post second vaccination had a somewhat higher odds of reporting symptoms regardless of Ct (any OR (also adjusted for age and sex)=1.35 (95% CI 1.05-1.73) p=0.02; classic OR=1.86 (95% CI 1.43-2.42) p<0.0001). There was no evidence of differential reporting of any symptoms for those 0-20 days post first vaccination (OR=1.20 (95% CI 0.82-1.74) p=0.34), but odds were modestly higher for classic symptoms after 14 June 2021 (OR=1.68 (95% CI 1.15-2.43) p=0.007). Again, reporting bias could contribute to some of these differences, given that the overall shape of the relationship between Ct and symptom reporting remained similar.

**Figure S7.**
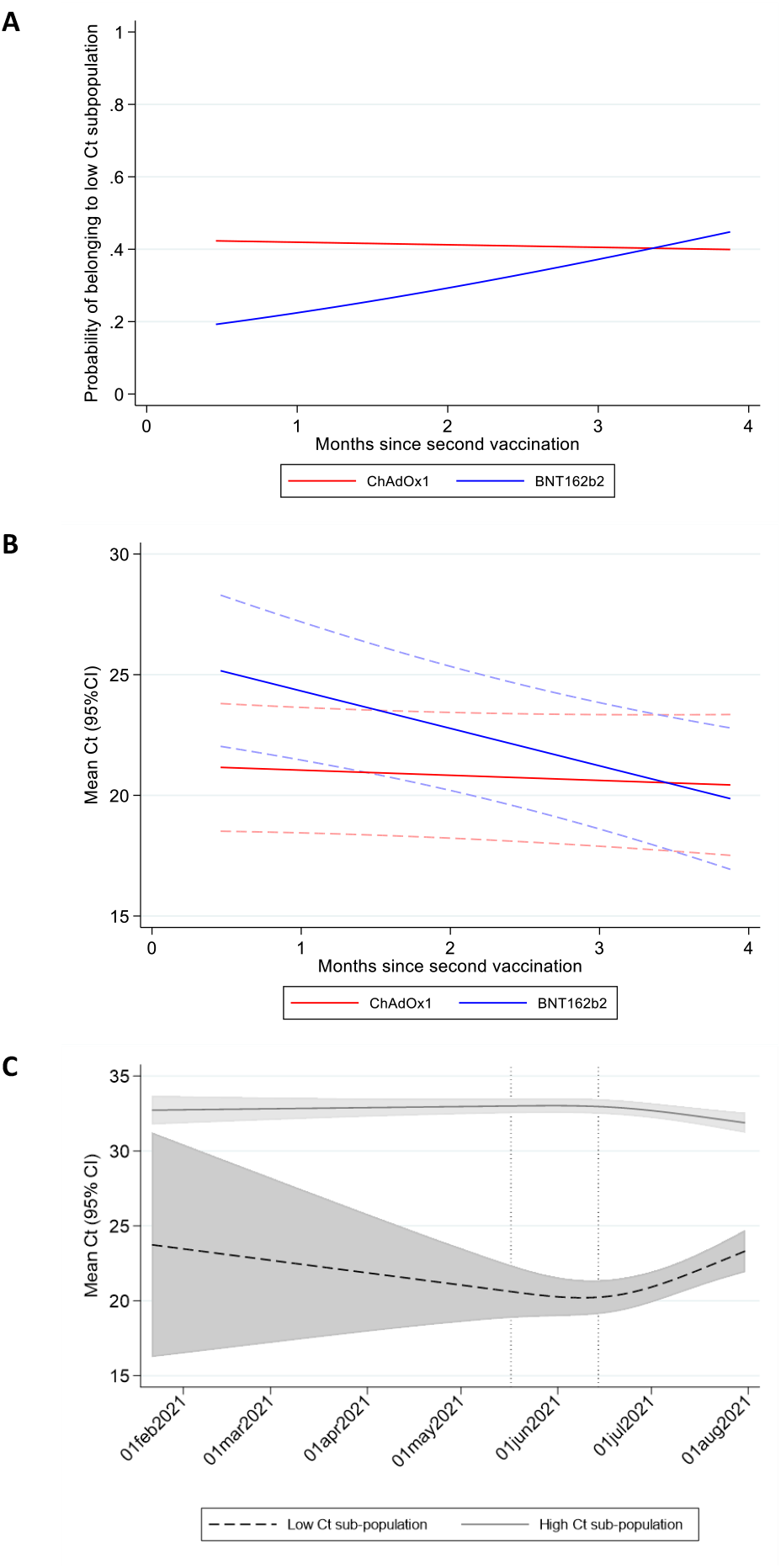
Adjusted effects of vaccine type and months since second vaccination on probability of belonging to the low Ct sub-population (A) and Ct values within the low Ct sub-population (B), and of calendar time on Ct values within the low Ct sub-population (C) in new PCR-positives ≥14 days after second vaccination in those aged ≥18 years. Note: estimated at the reference category for other factors (27 April 2021, male, no previous PCR/antibody-positive, not reporting a long-term health condition). In (C), test for non-linearity in effect of calendar date p=0.003 for low and <0.0001 for high Ct sub-population.

**Figure S8.**
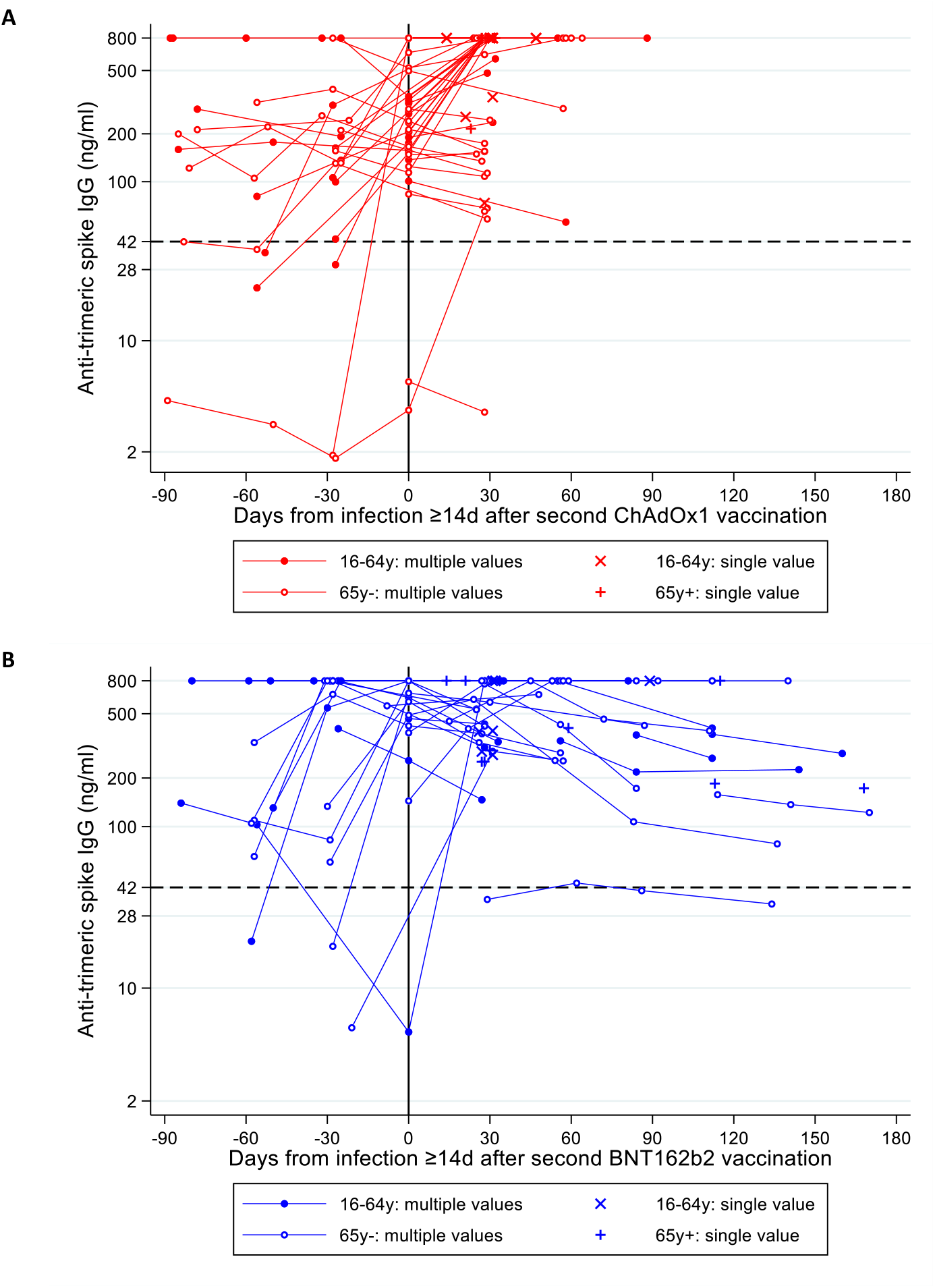
Antibody changes after new PCR-positive ≥14 days after second ChAdOx1 (N=60) (A) or BNT162b2 (N=51] (B) vaccinations. Median second vaccination date (IQR) 24 Apr 2021 (15 April-18 May) for ChAdOx1 and 5 April (9 January-16 April 2021) for BNT162b2. Median (IQR) new PCR-positive date 13 June (30 May-19 June) for ChAdOx1 and 25 May 2021 (20 February-16 June) for BNT162b2.

**Table S1:** Characteristics of visits included in the analyses. Note: analysis is based on visits rather than participants and restricted to those either being unvaccinated or vaccinated with ChAdOx1, BNT162b2 or mRNA-1273: factors above and vaccination exposure (detailed in **Table S2**) could change over time

| Characteristic | 1 Dec 2020-16 May 2021 (Alpha-dominant) | 17 May 2021 – 2 August (Delta-dominant) | 17 May 2021 – 2 August (Delta-dominant) | 17 May 2021 – 2 August (Delta-dominant) |
| --- | --- | --- | --- | --- |
|  | ≥18 years old | ≥18 years old | 18-34 years | 35-64 years |
|  | Total, n (%) or median (IQR) | Total, n (%) or median (IQR) | Total, n (%) or median (IQR) | Total, n (%) or median (IQR) |
| **Female** | 1,383,521 (53.6) | 439,890 (54.2) | 62,747 (54.6) | 234,514 (55.8) |
| **White ethnicity** | 2,418,424 (93.7) | 756,724 (93.2) | 101,134 (87.9) | 385,946 (91.9) |
| **Age** | 56 (41,68) | 57 (42,69) | 28 (23,32) | 52 (44,58) |
| **Region / country** |  |  |  |  |
| London | 451,476 (17.5) | 140,057 (17.3) | 29,154 (25.3) | 77,441 (18.4) |
| North West England | 290,953 (11.3) | 83,529 (10.3) | 11,435 (9.9) | 42,585 (10.1) |
| North East England | 95,547 (3.7) | 30,158 (3.7) | 3,892 (3.4) | 15,091 (3.6) |
| Yorkshire | 204,631 (7.9) | 69,546 (8.6) | 9,105 (7.9) | 36,058 (8.6) |
| West Midlands | 187,801 (7.3) | 62,202 (7.7) | 8,843 (7.7) | 31,657 (7.5) |
| East Midlands | 158,924 (6.2) | 50,893 (6.3) | 6,943 (6.0) | 26,228 (6.2) |
| South East England | 322,740 (12.5) | 106,483 (13.1) | 12,721 (11.1) | 55,197 (13.1) |
| South West England | 199,878 (7.7) | 56,290 (6.9) | 5,966 (5.2) | 27,392 (6.5) |
| East England | 259,811 (10.1) | 67,086 (8.3) | 7,974 (6.9) | 34,334 (8.2) |
| Northern Ireland | 73,296 (2.8) | 22,399 (2.8) | 3,146 (2.7) | 12,158 (2.9) |
| Scotland | 211,833 (8.2) | 73,518 (9.1) | 9,841 (8.6) | 38,064 (9.1) |
| Wales | 123,131 (4.8) | 49,463 (6.1) | 6,003 (5.2) | 23,914 (5.7) |
| **Household size** |  |  |  |  |
| 1 | 491,866 (19.1) | 168,423 (20.8) | 12,886 (11.2) | 74,749 (17.8) |
| 2 | 1,246,647 (48.3) | 388,494 (47.9) | 46,117 (40.1) | 166,359 (39.6) |
| 3 | 397,242 (15.4) | 122,254 (15.1) | 27,504 (23.9) | 80,720 (19.2) |
| 4 | 315,769 (12.2) | 95,518 (11.8) | 18,769 (16.3) | 73,227 (17.4) |
| 5+ | 128,497 (5.0) | 36,935 (4.6) | 9,747 (8.5) | 25,064 (6.0) |
| **Multigenerational household** | 111,165 (4.3) | 35,389 (4.4) | 4,825 (4.2) | 28,576 (6.8) |
| **Rural-urban classification** |  |  |  |  |
| Major urban | 915,065 (35.5) | 292,442 (36) | 53,528 (46.5) | 156,941 (37.4) |
| Urban city/town | 1,095,963 (42.5) | 344,593 (42.5) | 45,766 (39.8) | 177,297 (42.4) |
| Rural town | 279,076 (10.8) | 86,148 (10.6) | 8,470 (7.4) | 42,239 (10.1) |
| Rural village | 289,917 (11.2) | 88,441 (10.9) | 7,259 (6.3) | 43,642 (10.4) |
| **Deprivation centile (1 = most deprived, 10 = least deprived)** | 6 (3,8) | 6 (3,8) | 5 (3,7) | 6 (3,8) |
| **Ever reported to be a care home worker** | 30,050 (1.2) | 9,266 (1.1) | 1,742 (1.5) | 6,739 (1.6) |
| **Ever reported to be a person-facing healthcare worker** | 66,238 (2.6) | 20,850 (2.6) | 4,134 (3.6) | 15,219 (3.6) |
| **Ever reported to be person-facing social care worker** | 29,691 (1.2) | 10,261 (1.3) | 1,705 (1.5) | 7,687 (1.8) |
|  | ≥18 years old | ≥18 years old | 18-34 years | 35-64 years |
|  | Total, n (%) or median (IQR) | Total, n (%) or median (IQR) | Total, n (%) or median (IQR) | Total, n (%) or median (IQR) |
| **Ever reported to have a long-term health condition** | 722,819 (28.0) | 231,416 (28.5) | 20,099 (17.5) | 101,659 (24.2) |
| **Days since previous visit** |  |  |  |  |
| >14 days | 1,772,532 (68.7) | 707,099 (87.1) | 97,880 (85.1) | 367,294 (87.4) |
| ≤ 14 days | 662,234 (25.7) | 78,135 (9.6) | 12,671 (11.0) | 39,603 (9.4) |
| Enrolment | 145,255 (5.6) | 26,390 (3.3) | 4,472 (3.9) | 13,222 (3.1) |
| **Smoking status** |  |  |  |  |
| Non-smoker | 2,325,890 (90.2) | 731,654 (90.1) | 101,670 (88.4) | 370,618 (88.2) |
| Tobacco smoker | 195,301 (7.6) | 61,758 (7.6) | 9,915 (8.6) | 37,509 (8.9) |
| Only vape | 58,830 (2.3) | 18,212 (2.2) | 3,438 (3.0) | 11,992 (2.9) |
| **Contact with hospital in last 28 days** |  |  |  |  |
| Missing (n) | 34,645 | 2,607 | 369 | 1,320 |
| No | 1,995,000 (78.4) | 604,317 (74.7) | 84,146 (73.4) | 312,112 (74.5) |
| Yes, I have | 347,145 (13.6) | 138,498 (17.1) | 17,817 (15.5) | 71,921 (17.2) |
| No, but someone in household has | 203,231 (8.0) | 66,202 (8.2) | 12,691 (11.1) | 34,766 (8.3) |
| **Contact with care home in last 28 days** |  |  |  |  |
| Missing (n) | 42,063 | 3,628 | 527 | 1,821 |
| No | 2,452,588 (96.6) | 766,973 (94.9) | 108,837 (95.1) | 394,263 (94.3) |
| Yes, I have | 49,649 (2.0) | 26,407 (3.3) | 2,722 (2.4) | 15,791 (3.8) |
| No, but someone in household has | 35,721 (1.4) | 14,616 (1.8) | 2,937 (2.6) | 8,244 (2.0) |

**Table S2:** Vaccination and previous infection status for visits included in analysis (18 years and older) Note: analysis is based on visits rather than participants: factors (**Table S1**) and vaccination exposure could change over time. See methods for definition of PCR-positive episodes of infection and prior positivity. Cells with zeros only are by definition. NA indicates to few visits to estimate effectiveness.

|  | **1 Dec 2020 – 16 May 2021 (Alpha-dominant)** |  | **17 May 2021 – 2 August (Delta-dominant)** |  |
| --- | --- | --- | --- | --- |
| **Vaccination and previous infection status** | **No prior study or national testing programme swab-positive and no antibody positive**<br>**Number of visits (row %)**<br>**[number of positives]** | **Prior study or national testing programme swab-positive or antibody positive**<br>**Number of visits (row %)**<br>**[number of positives]** | **No prior study or national testing programme swab-positive and no antibody positive**<br>**Number of visits (row %)**<br>**[number of positives]** | **Prior study or national testing programme swab-positive or antibody positive**<br>**Number of visits (row %)**<br>**[number of positives]** |
| Not vaccinated, no prior positive, >21 days before vaccination | 1,561,154 (100%) [14,440] | 0 (0%) [0] | 27,135 (100%) [351] | 0 (0%) [0] |
| Not vaccinated, no prior positive, 1-21 days before vaccination | 220,749 (100%) [665] | 0 (0%) [0] | 16,754 (100%) [51] | 0 (0%) [0] |
| Vaccinated 0-20 days ago, ChAdOx1 | 127,572 (95%) [293] | 4,169 (5%) [11] | 4,435 (92%) [4] | 387 (8%) [0] |
| Vaccinated 0-20 days ago, BNT162b2 | 70,837 (98%) [419] | 1,562 (2%) [6] | 23,387 (90%) [175] | 2,496 (10%) [10] |
| Vaccinated 0-20 days ago, mRNA-1273 | NA | NA | 2,941 (91%) [19] | 275 (9%) [1] |
| ≥ 21 days after 1<sup>st</sup> dose, no second dose ChAdOx1 | 236,306 (95%) [227] | 11,390 (5%) [11] | 64,232 (91%) [194] | 6,377 (9%) [20] |
| ≥ 21 days after 1<sup>st</sup> dose, no second dose, BNT162b2 | 141,968 (96%) [278] | 5,259 (4%) [11] | 38,505 (91%) [410] | 3,998 (9%) [12] |
| ≥ 21 days after 1<sup>st</sup> dose, no second dose, mRNA-1273 | NA | NA | 5,710 (91%) [30] | 534 (9%) [2] |
| 0-13 days after second dose ChAdOx1 | 32,239 (95%) [14] | 1,657 (5%) [2] | 44,879 (91%) [91] | 4,429 (9%) [1] |
| 0-13 days after second dose, BNT162b2 | 34,406 (96%) [28] | 1,542 (4%) [0] | 11,146 (90%) [40] | 1,184 (10%) [1] |
| ≥ 14 days after second dose ChAdOx1 | 30,178 (95%) [9] | 1,562 (5%) [1] | 303,511 (92%) [1,175] | 27,166 (8%) [61] |
| ≥ 14 days after second dose, BNT162b2 | 70,058 (96%) [39] | 3,231 (4%) [4] | 199,411 (93%) [422] | 16,166 (7%) [25] |
| Not vaccinated, prior positive | 0 (0%) [0] | 24,256 (100%) [80] | 0 (0%) [0] | 5,753 (100%) [25] |

**Table S3:** Vaccine effectiveness by subgroups during the Delta-dominant period in those aged 18-64 years. Note: obtained using a model with logit-linear slopes on time since ≥14 days after second ChAdOx1 or BNT162b2 vaccination. The effect estimates in the table relate to the effectiveness on the 14^th^ day after the second dose (‘intercept’ of the second dose effect). Potential waning of effectiveness as a function of time since ≥14 days after the second vaccination is obtained from the same model by including logit-linear time terms for BNT162b2 and ChAdOx1 that are not varied by subgroup due to insufficient numbers to estimate different slopes by the combination of vaccine type and subgroup effects (potential waning effects shown in **Figure 2, Figure S3** & **Figure S4**). P-values<0.01 shown in bold (0.01 threshold used given the number of tests performed).

|  | VE against all infections |  |  | VE against infections with Ct<30 |  |  | VE against infections with reported symptoms |  |  |
| --- | --- | --- | --- | --- | --- | --- | --- | --- | --- |
| By evidence of prior infection | No evidence of prior infection | Evidence of prior infection | Heterogeneity p-value | No evidence of prior infection | Evidence of prior infection | Heterogeneity p-value | No evidence of prior infection | Evidence of prior infection | Heterogeneity p-value |
| $\geq 21$ days after 1st dose, no second vaccination BNT162b2 | 57% (50-63%) | 91% (84-95%) | **<0.0001** | 62% (54-68%) | 96% (88-98%) | **<0.0001** | 58% (50-65%) | 94% (87-98%) | **<0.0001** |
| $\geq 21$ days after 1st dose, no second vaccination ChAdOx1 | 43% (30-54%) | 53% (24-71%) | 0.45 | 45% (30-57%) | 78% (49-90%) | 0.03 | 34% (16-48%) | 83% (55-94%) | **0.007** |
| $\geq 21$ days after 1st dose, no second vaccination mRNA-1273 | 76% (64-83%) | 86% (44-97%) | 0.43 | 76% (62-85%) | 100% (100-100%) | **<0.0001** | 76% (62-85%) | 100% (100-100%) | **<0.0001** |
| 14 days after 2nd dose BNT162b2 | 85% (79-90%) | 93% (87-96%) | **0.006** | 92% (87-95%) | 98% (94-99%) | **0.004** | 93% (89-97%) | 99% (96-100%) | **0.002** |
| 14 days after 2nd dose ChAdOx1 | 68% (61-73%) | 88% (83-92%) | **<0.0001** | 69% (61-75%) | 92% (87-95%) | **<0.0001** | 72% (64-78%) | 94% (89-97%) | **<0.0001** |
| By age | Aged 18-34 years old | Aged 35-64 years old | Heterogeneity p-value | Aged 18-34 years old | Aged 35-64 years old | Heterogeneity p-value | Aged 18-34 years old | Aged 35-64 years old | Heterogeneity p-value |
| $\geq 21$ days after 1st dose, no second vaccination BNT162b2 | 64% (57-70%) | 36% (14-53%) | **0.0009** | 68% (60-74%) | 46% (22-62%) | **0.01** | 66% (58-72%) | 32% (0-53%) | 0.002 |
| $\geq 21$ days after 1st dose, no second vaccination ChAdOx1 | 43% (11-64%) | 18% (-9-39%) | 0.18 | 57% (21-77%) | 22% (-9-45%) | 0.10 | 37% (-7% -63%) | 0% (-43-31%) | 0.16 |
| ≥21 days after 1st dose, no second vaccination mRNA-1273 | 82% (70-89%) | 56% (23-75%) | 0.02 | 81% (66-90%) | 64% (27-83%) | 0.18 | 85% (72-92%) | 48% (-1-73%) | **0.008** |
| 14 days after 2nd dose BNT162b2 | 90% (85-93%) | 77% (65-85%) | **0.0001** | 95% (91-97%) | 88% (79-93%) | **0.002** | 96% (93-98%) | 88% (78-94%) | **<0.0001** |
| 14 days after 2nd dose ChAdOx1 | 73% (65-80%) | 54% (40-65%) | **0.002** | 74% (64-81%) | 57% (41-69%) | 0.02 | 76% (67-83%) | 57% (39-70%) | **0.007** |
| **By interval between first and second doses** | **Dosing interval <9 weeks** | **Dosing interval ≥9 weeks** | **Heterogeneity p-value** | **Dosing interval <9 weeks** | **Dosing interval ≥9 weeks** | **Heterogeneity p-value** | **Dosing interval <9 weeks** | **Dosing interval ≥9 weeks** | **Heterogeneity p-value** |
| 14 days after 2nd dose BNT162b2 | 85% (78-90%) | 85% (78-90%) | 0.89 | 93% (88-96%) | 92% (86-95%) | 0.49 | 93% (88-96%) | 93% (88-96%) | 0.97 |
| 14 days after 2nd dose ChAdOx1 | 68% (60-75%) | 67% (61-73%) | 0.80 | 69% (60-77%) | 69% (61-75%) | 0.89 | 74% (65-80%) | 71% (64-78%) | 0.47 |
| **By self-reported long-term health conditions** | **No long-term health conditions** | **Long-term health conditions** | **Heterogeneity p-value** | **No long-term health conditions** | **Long-term health conditions** | **Heterogeneity p-value** | **No long-term health conditions** | **Long-term health conditions** | **Heterogeneity p-value** |
| ≥21 days after 1st dose, no second vaccination BNT162b2 | 58% (50-64%) | 62% (39-77%) | 0.65 | 62% (54-69%) | 66% (40-80%) | 0.76 | 58% (49-66%) | 66% (40-81%) | 0.48 |
| ≥21 days after 1st dose, no second vaccination ChAdOx1 | 45% (32-55%) | 7% (-48-41%) | 0.04 | 46% (30-58%) | 28% (-27-59%) | 0.35 | 38% (19-52%) | 5% (-66-46%) | 0.18 |
| ≥21 days after 1st dose, no second vaccination mRNA-1273 | 77% (65-84%) | 59% (-17-85%) | 0.31 | 78% (65-87%) | 59% (-35-88%) | 0.33 | 80% (66-88%) | 44 (-61-81%) | 0.09 |
| 14 days after 2nd dose BNT162b2 | 86% (80-90%) | 81% (69-89%) | 0.23 | 92% (87-95%) | 92% (85-96%) | 0.96 | 94% (89-96%) | 92% (84-96%) | 0.38 |
| 14 days after 2nd dose ChAdOx1 | 69% (62-74%) | 58% (39-71%) | 0.10 | 70% (62-76%) | 65% (46-77%) | 0.48 | 73% (65-79%) | 64% (44-77%) | 0.23 |

**Table S4:** Independent associations with Ct values in new PCR-positives ≥14 days after second vaccination 18 years and older.

|  | Low Ct vs high Ct:<br>odds of class membership |  |  | Low Ct sub-population: mean Ct |  |  | High Ct sub-population: mean Ct |  |  | Het low vs high mean |
| --- | --- | --- | --- | --- | --- | --- | --- | --- | --- | --- |
|  | OR | 95% CI | p | Effect | 95% CI | p | Effect | 95% CI | p | p |
| Overall mean Ct |  |  |  | **+21.2** | **+18.5,+23.8** | **<0.0001** | **33.0** | **+32.4,+33.5** | **<0.0001** | **<0.0001** |
| Per month post 1 Jan 2021 (reference 27 April) 2021 | **1.76** | **1.41,2.20** | **<0.0001** | **Fig S5C** |  | **0.0002** | **Fig S5C** |  | **0.0004** |  |
| Vaccine: 14 days post second dose |  |  |  |  |  |  |  |  |  |  |
| ChAdOx1 | 1.00 (ref) |  | - | 0.0 (ref) |  | - | 0.0 (ref) |  | - |  |
| BNT162b2 | **0.33** | **0.16,0.67** | **0.002** | **+4.0** | **+1.4,+6.6** | **0.003** | +0.5 | -0.2,+1.2 | 0.16 | 0.008 |
| Vaccine: per month later post second dose |  |  |  |  |  |  |  |  |  |  |
| ChAdOx1 | 0.97 | 0.79,1.19 | 0.78 | -0.2 | -0.8,+0.3 | 0.45 | -0.1 | -0.3,+0.2 | 0.66 | 0.58 |
| BNT162b2 | **1.43** | **1.07,1.91** | **0.01** | **-1.6** | **-2.5,-0.6** | **0.002** | -0.0 | -0.3,+0.3 | 0.82 | 0.01 |
| Previously PCR/antibody-positive | **0.35** | **0.21,0.57** | **<0.0001** |  |  | *0.58* |  |  | *0.10* |  |
| Report having a long-term health condition | **0.69** | **0.53,0.90** | **0.006** |  |  | *0.78* |  |  | *0.52* |  |
| Sex: Male |  |  | *0.34* | 0.0 (ref) |  | - | 0.0 (ref) |  | - |  |
| Female |  |  | - | +0.0 | -0.7,+0.7 | 0.98 | **+0.3** | **+0.0,+0.6** | **0.04** | 0.39 |
| Age |  |  | *0.19* |  |  | *0.24* |  |  | *0.64* |  |
| Ethnicity (white vs non-white) |  |  | *0.96* |  |  | *0.65* |  |  | *0.85* |  |
| Patient facing healthcare worker |  |  | *0.63* |  |  | *0.90* |  |  | *0.28* |  |
| Deprivation percentile |  |  | *0.60* |  |  | *0.94* |  |  | *0.91* |  |
| Interval between first and second dose |  |  | *0.83* |  |  | *0.64* |  |  | *0.16* |  |

Note: where effect size not shown (gray cells), p-values in italics show the additional effect of adding this term into the model for either class membership or for and effect on mean Ct in the two sub-populations. Test for interaction between effect of vaccine type and time since second dose: p=0.02 for membership of high vs low Ct sub-population, p=0.01 for low mean Ct, p=0.89 for high mean Ct. Excluding 17 positives occurring $\geq 14$ days after second vaccination but not reporting either two ChAdOx1 or two BNT162b2 doses. There was no evidence of interaction between vaccine type and age for class probability (p=0.09) or mean Ct (p>0.1); nor was there evidence of interaction between vaccine type and date for class probability (p=0.69) or mean Ct (p>0.4).

